# SPINAL CORD STIMULATION IMPROVES MOTOR FUNCTION AND SPASTICITY IN CHRONIC POST-STROKE UPPER LIMB HEMIPARESIS

**DOI:** 10.1101/2025.08.01.25332445

**Authors:** Roberto M. de Freitas, Shovan Bhatia, Erynn Sorensen, Nikhil Verma, Erick Carranza, Scott Ensel, Luigi Borda, Amy Boos, Jeff Goldsmith, Lee E. Fisher, Daryl P. Fields, Marc P. Powell, Shane Gordon, Jeffrey Balzer, Robert M. Friedlander, George F. Wittenberg, Peter Gerszten, John W. Krakauer, Elvira Pirondini, Douglas J. Weber, Marco Capogrosso

## Abstract

Here, we report the final outcomes of a pilot clinical trial testing preliminary efficacy and safety of cervical epidural spinal cord stimulation (SCS) for chronic post-stroke upper-limb hemiparesis (NCT04512690). We implanted seven participants with profound motor deficits (Fugl-Meyer Assessment [FMA] scores 15–35) using two leads implanted unilaterally in the cervical spinal cord for 4 weeks. Under SCS ON, motor function immediately improved regardless of impairment severity (average +32% strength and +5.6 FMA-points). Notably, 3/7 participants with residual corticospinal connectivity to finger muscles regained hand/finger function with SCS. Despite performing only 8.6hrs of motor activity (5.5hrs with SCS ON), participants improved by average +6.6 FMA-points at the end of the study compared to baseline and spasticity decreased in all participants. While all benefited, our preliminary analysis indicates that spared sensory function may be a determinant of responsiveness to SCS. No serious adverse events occurred.

## INTRODUCTION

Stroke is the leading cause of arm paralysis in the U.S.A^1,2^. Every year, approximately 800,000 individuals suffer from cerebral stroke, of whom 400,000 experience chronic arm paresis^3–5^. Because of the significant impact that stroke has on their quality of life, affected individuals rank the recovery of arm and hand function as their first unmet clinical priority^6^. After stroke, the loss of corticospinal axons leads to a stereotypical manifestation of chronic hemiparesis^7,8^, characterized by: loss of strength, *i.e.,* loss of the ability to effectively activate muscles and generate forces; loss of dexterity, *i.e.,* loss of the ability to grade force and control digits; spasticity, *i.e.*, velocity-dependent resistance to passive stretch of limb joints^9,10^; and intrusion of abnormal synergies, *i.e.,* abnormal co-contraction of muscles during voluntary movements.

Despite decades of research indicating that high-dose motor rehabilitation (100-120 hours in 4 weeks) can improve these symptoms^11,12^, the rehabilitation dose in actual clinical practice remains substantially lower (∼7 hours in 4 weeks), falling well short of these goals^13^. As a result, significant research efforts have historically been directed towards the development of neuromodulation technologies that could catalyze the effects of rehabilitation^14–16^. That is, these technologies aim to induce lasting therapeutic effects by modulating the nervous system activity during motor training, resulting in post-intervention improvements in absence of stimulation. Most notably, vagus-nerve stimulation (VNS) was recently approved by the U.S.A. Food and Drug Administration (FDA) for use in people with post-stroke hemiparesis^14^. When combined with 27 hours of motor rehabilitation, VNS increased Fugl-Meyer Assessment (FMA) scores for upper extremity motor function more than rehabilitation alone.

In contrast to these approaches, we sought to design an intervention that could offer immediate and assistive improvements in motor function without the need for formal motor rehabilitation. Recent studies suggested that epidural spinal cord stimulation (SCS) may improve strength^17^, dexterity^17^, spasticity^18,19^, and synergies^20^ in people with spinal cord injury and other motor disorders. Cervical SCS selectively activates dorsal root sensory afferents that project to upper-limb motor neuron pools^21–23^. Through this excitatory monosynaptic pathway, SCS can immediately facilitate voluntary movements in the upper limbs even without rehabilitation^24,25^. In addition to these assistive effects (*i.e.,* changes in motor function that occur immediately as SCS is turned ON), SCS can also enable therapeutic effects over time, resulting in improvements of motor function in absence of stimulation)^26,27^. Therefore, different from other neuromodulation technologies^14,15^, SCS uniquely combines these two interdependent effects to drive *effective improvements*^24,26^. Following this rationale, we previously reported preliminary results from the first two study participants of a first-in-human clinical trial (NCT04512690) to prospectively characterize the assistive and therapeutic effects of epidural cervical SCS in participants with chronic post-stroke hemiparesis^24^. Previously, we observed larger assistive effects in one participant with milder impairments, but significantly smaller improvements in the other with more severe impairments. This contrast raised questions about the reproducibility of the assistive effects of SCS across participants. Additionally, because the emergence of therapeutic effects was unanticipated, we initially overlooked how they might combine with assistive effects over time to drive effective improvements. This left critical questions on the feasibility of SCS for post-stroke hemiparesis unanswered: how reproducible are the assistive effects of SCS across the variability of the post-stroke phenotype observed in clinical settings? Do baseline motor impairments determine the degree of responsiveness to SCS? Do assistive effects change over time? What is the maximum effective improvement that can be achieved? Answering these questions is crucial to advance SCS in the path towards FDA approval and clinical use.

Here, we report the final results of our first-in-human clinical trial (NCT04512690) in *n* = 7 participants with chronic post-stroke hemiparesis. Participants were implanted with two percutaneous leads in the cervical spinal cord for a period of 4 weeks during which they were assessed on a daily basis with a battery of motor tasks. Importantly, participants did not participate in a formal rehabilitation protocol during the 4-week period. After 4 weeks, the leads were explanted. Participants included a broad spectrum of post-stroke impairments ranging from very severe (15 FMA) to moderate (35 FMA) and including both ischemic and hemorrhagic strokes. By characterizing both the magnitude and variability of SCS effect sizes, as well as exploring potential determinants of SCS responsiveness (e.g., integrity of corticospinal tract and sensory impairment), we assessed safety and preliminary efficacy of SCS for post-stroke chronic hemiparesis.

## RESULTS

We conducted a non-randomized clinical pilot study (NCT04512690) aimed at testing the efficacy and safety of epidural cervical SCS in *n* = 7 individuals affected with chronic hemiparesis after stroke (**Fig. 1**). A list of all outcomes of this clinical trial, including those analyzed in this study, is provided in **Supplementary Information Table 1**. We assessed the efficacy of cervical SCS by measuring: (*i*) assistive effects immediately enabled in motor function when SCS was delivered, *i.e*., the difference between SCS ON and SCS OFF conditions; (*ii*) therapeutic effects emerged over time in the absence of SCS, *i.e.*, the difference between timepoints under the SCS OFF condition ; as well as (*iii*) effective improvement, which represents the total functional gain achievable with SCS as a neuroprosthetic device, *i.e.*, the difference between pre-implant baseline and SCS ON condition (**Fig. 1a**). The study was approved by the University of Pittsburgh Institutional Review Board (IRB STUDY19090210).

**Fig. 1.**
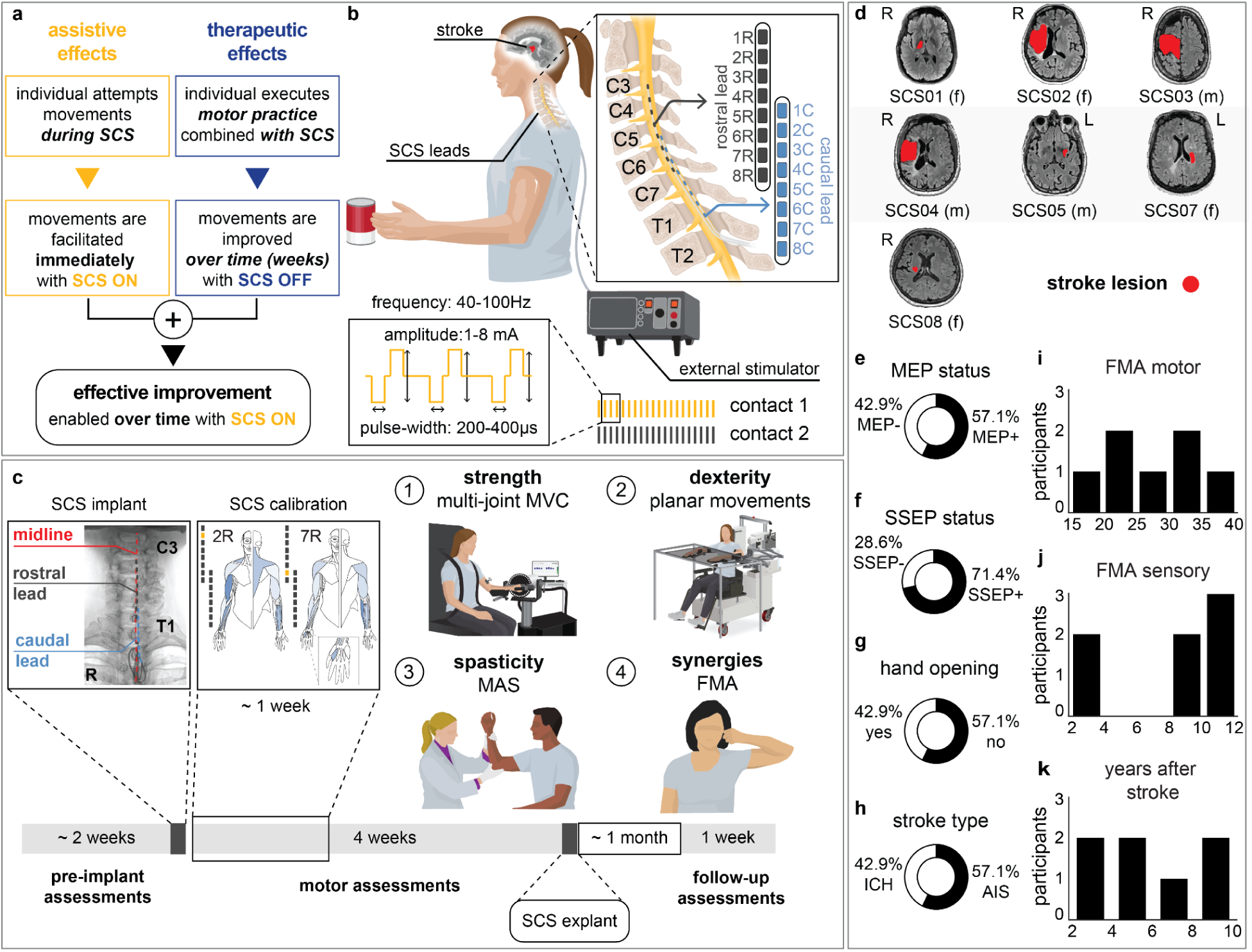
Study design and participant cohort. **a,b.** Study design and overview. **a.** Conceptual definition of the effects enabled by spinal cord stimulation (SCS) on motor function: *assistive effects* refer to immediate motor improvements observed with stimulation ON; *therapeutic effects* are improvements in motor function over time with stimulation OFF; *effective improvement* reflects the combination of both assistive and therapeutic effects, and is assessed as the change in motor function from the pre-implant baseline to a given timepoint with stimulation ON. **b.** Illustration of cervical SCS applied using two 8-contact epidural leads. Stimulation was administered via an external multi-channel stimulator. **c.** Clinical trial timeline: pre-implant baseline assessments, surgical implantation, calibration of SCS parameters, and motor assessments. Motor assessments included: (1) strength (multi-joint maximum voluntary contraction, MVC, force), (2) dexterity (planar reach-and-pull kinematics), (3) spasticity (Modified Ashworth Scale, MAS), and (4) synergies (subcomponents Fugl-Meyer Assessment, FMA). **d-k.** Participant cohort and baseline characteristics. **d.** Stroke lesions (red) across participants annotated on T1-weighted Fluid Attenuated Inversion Recovery Magnetic Resonance Imaging (FLAIR MRI). **e.** Motor evoked potential (MEP) status assessed with transcranial magnetic stimulation, **f.** Somatosensory evoked potential (SSEP) status assessed with ulnar nerve stimulation, **g.** Hand opening ability, **h.** Stroke type (ICH, intracerebral hemorrhage, vs. AIS, acute ischemic stroke). **i.** Baseline FMA motor scores, **j.** Baseline FMA sensory scores, **k.** Years after stroke.

### Study design and experimental setup

Participants were surgically implanted with a pair of 8-contact SCS leads (PN 977A260, Medtronic) in the dorsal epidural space spanning from C3 to T1 spinal levels, where sensory afferents and motoneurons innervating the upper limb muscles are located (**Fig. 1b; Extended Data Fig. 1**). Using intraoperative mapping^24^, both electrodes were positioned lateral to the spinal midline, such that large-diameter sensory fibers running in the dorsal roots ipsilateral to the paretic limb were preferentially activated (**Extended Data Fig. 1a-g**)^23,25,28^. Over a period of 4 weeks, after a calibration step to identify functional stimulation parameters **(Fig. 1c; Extended Data Fig. 2**), we conducted motor assessments with and without SCS for quantifying: strength, measured as the maximum voluntary contraction (MVC) force produced during arm and hand movements; dexterity, quantified through arm kinematics during a planar reaching task; spasticity, quantified using the Modified Ashworth Scale (MAS) for different upper limb joints; and synergies, primarily measured using the Fugl-Meyer Assessment (FMA) motor scores for the upper extremity (**Fig. 1c**). Importantly, we did not implement a formal motor rehabilitation protocol. Instead, participants were involved 5 days a week for about 4 hours in scientific and motor assessments for a total of 19 experimental sessions. Despite the assessments are not formally an exercise protocol, when discarding for set-up, idle time and tests, such as impedance checks and patient care, we measured that participants effectively engaged in active movement during assessments for a total of 8.6 hours of “motor activities” over the 4 weeks of intervention (on average 5.5 hours with SCS ON plus 3.1 hours with SCS OFF, **Supplementary Information Table 2**). SCS was administered using a custom stimulation system^24^, which was used only while the participant was in the laboratory (**Supplementary Information Methods**). As the primary outcome of the study, safety was assessed by systematically logging all adverse events. The lead electrodes were surgically removed from all participants after the 4-week testing period. Participants returned for follow-up clinical assessments one month after lead explantation (follow-up outcomes in **Supplementary Information Tables 3-5**).

### Participants

We enrolled a total of *n* = 8 participants, however only *n* = 7 completed the study (SCS06 withdrew due to an incipient health condition prior to implant). All participants presented with marked chronic hemiparesis symptoms resulting from brain lesions caused by hemorrhagic or ischemic stroke (**Fig. 1d; Extended Data Table 1**). Notably, a broad spectrum of motor and sensory impairments was observed across participants (**Fig. 1e-k; Extended Data Table 1**). Aligned with our study aim, our inclusion and exclusion criteria were defined to enable the enrollment of a representative range of hemiparesis symptoms (**see Methods**).

### Safety

To assess safety of epidural cervical SCS applied to individuals with chronic post-stroke hemiparesis, we systematically documented all adverse events (AEs) throughout the clinical trial. All documented adverse events (AEs) are described in **Table 1**. There were 14 mild AEs in this clinical trial, i.e., no moderate or severe AEs were observed. All AEs were resolved rapidly without sequelae. Most AEs (11/14) did not have a definite and direct relationship with the study, except for three AEs with participants SCS01, SCS04, and SCS05 (**Extended Data Table 2**). In particular, SCS04 experienced shortness of breath when stimulation was delivered at high amplitudes (5 mA) using a rostral electrode located between C3 and C4 spinal levels. This adverse event was resolved immediately by discontinuing stimulation. We believe this undesired side effect resulted from stimulation of the phrenic nerve in the C3/C4 spinal levels, likely interfering with diaphragm function^29^. In response, we reported this AE to the IRB and FDA and modified our protocol to avoid stimulation of the most rostral cervical spinal levels as a safety precaution. Specifically, stimulation of rostral contacts (commonly 1R - 3R) at high amplitudes (above 3 mA) was avoided. Due to the rostro-caudal organization of motor neuron pools in the cervical spinal cord^30^, this protocol modification limited the ability to recruit proximal muscles, such as trapezius and deltoid muscles.

**Table 1.** Adverse events. Summary of all adverse events documented throughout the clinical trial for each participant. Adverse events were recorded using a logging form provided by the Institutional Review Board (IRB) of the University of Pittsburgh.

| ID | number of events | severity |  |  | relationship with the study |  |  | serious events |
| --- | --- | --- | --- | --- | --- | --- | --- | --- |
|  |  | mild | moderate | severe | not related | possibly related | related |  |
| SCS01 | 2 | 2 | 0 | 0 | 0 | 1 | 1 | 0 |
| SCS02 | 1 | 1 | 0 | 0 | 0 | 1 | 0 | 0 |
| SCS03 | 0 | 0 | 0 | 0 | 0 | 0 | 0 | 0 |
| SCS04 | 3 | 3 | 0 | 0 | 0 | 2 | 1 | 0 |
| SCS05 | 2 | 2 | 0 | 0 | 0 | 1 | 1 | 0 |
| SCS07 | 6 | 6 | 0 | 0 | 2 | 4 | 0 | 0 |
| SCS08 | 0 | 0 | 0 | 0 | 0 | 0 | 0 | 0 |
| **TOTAL** | **14** | **14** | **0** | **0** | **2** | **9** | **3** | **0** |

### Systematic calibration of SCS parameters

After implantation, we conducted a systematic procedure to find stimulation parameters that immediately facilitated voluntary movements across multiple joints (**Fig. 2a,b; Extended Data Fig. 2a-c**). First we identified contacts that selectively targeted muscles involved in specific movements (e.g., biceps brachii for elbow flexion) using recruitment curves obtained with single-pulse stimulation with increasing amplitude (1 Hz, biphasic 200 µs square pulses, 0.1-10 mA). We then adjusted stimulation amplitude and frequency by observing immediate changes in movement range-of-motion, strength and speed when SCS was turned ON. Stimulation delivered at 40 Hz with amplitudes that evoked paresthesia often resulted in optimal assistive effects.

**Fig. 2.**
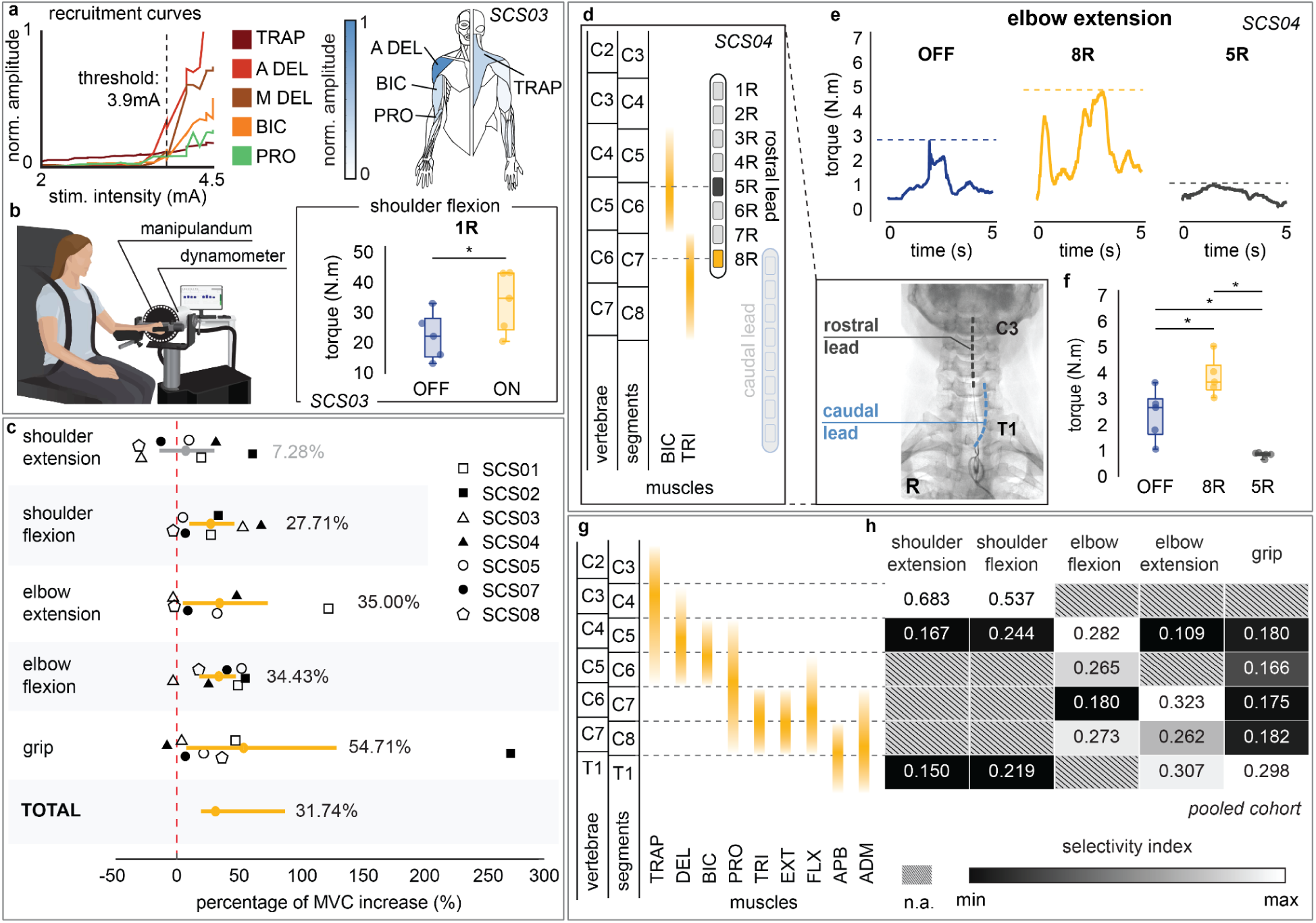
Assistive effects of spinal cord stimulation (SCS) on strength. **a,b.** SCS selectively activated shoulder muscles, enabling improved shoulder flexion strength in the SCS03 participant. **a.** Example recruitment curve from participant SCS03, showing selective activation of the proximal shoulder muscles at a threshold of 3.9 mA. Abbreviations: TRAP, trapezius; A DEL, anterior deltoid; M DEL, medial deltoid; BIC, biceps brachii; PRO, pronator. The human drawing illustrates normalized peak-to-peak electromyography (EMG) responses across muscles, with anterior (left) and posterior (right) views. **b.** Shoulder torque (N.m) was measured during isometric contractions using the isokinetic dynamometer (HUMAC Norm), illustrated on the left. SCS enabled immediate improvement in shoulder flexion strength for SCS03 when 1R contact targeting shoulder muscles was stimulated, i.e., significant assistive effects. **c.** Assistive effects of SCS on maximum voluntary contraction (MVC) force pooled across all participants. We observed significant improvements in strength across all isolated movements except for shoulder extension. On average, SCS enabled increased MVC strength by 32% across isolated movements. **d-f.** SCS selectivity was critical to improve MVC strength during elbow extension in the SCS04 participant. **d.** X-ray and spinal segment map illustrating the lead placement in SCS04 participant: contact 5R targeting C6 (biceps brachii) and contact 8R targeting C7 (triceps brachii). **e-f.** Example of elbow extension MVC task in SCS04 showing increased torque with stimulation at 8R (triceps) and decreased torque when stimulating at 5R (biceps). Horizontal bars with asterisks indicate statistically significant differences in **(f)**. **g,h.** Spinal segment map and selectivity index pooled across all participants. **g.** Pooled spinal segment map across participants obtained from intraoperative neurophysiological data. Abbreviations: TRI, triceps brachii; EXT, wrist extensors; FLX, wrist flexors; APB, abductor pollicis brevis; ADM, abductor digiti minimi. **h.** Selectivity index associated with significant increases in MVC strength. The heatmap shows that selective activation of appropriate spinal levels was crucial to enable assistive effects of SCS on strength.

We then fine-tuned these contacts together, balancing their stimulation amplitudes to target participant-specific impairments while minimizing interference between their effects on motor function (**Extended Data Fig. 2c, see Methods)**. For instance, in participants with pronounced flexor synergies, stimulation of contacts targeting elbow flexors (e.g., biceps brachii) could exacerbate abnormal muscle coactivation, thereby hindering elbow extension. These patient-specific factors were taken into account when calibrating SCS parameters using multiple contacts.

### Assistive effects of SCS on strength

Considering that SCS directly amplifies supraspinal inputs through an excitatory spinal pathway, we expected an immediate increase of strength when SCS was turned ON. To assess these assistive effects, we compared the torque produced during isometric maximum voluntary contraction (MVC) with SCS ON versus OFF conditions. Specifically, MVC was measured during shoulder flexion, shoulder extension, elbow flexion and elbow extension using an isokinetic dynamometer (**Fig. 2b**, HUMAC Norm.), whereas grip force was measured using a hydraulic hand-held dynamometer **(see Methods)**. On average, MVC strength increased significantly during shoulder flexion (28%), elbow extension (35%), elbow flexion (34%) and grip (55%). Overall, the mean effect of SCS on MVC strength across joints was estimated as an increase of 32% (**Fig. 2c)**. Importantly, despite inter-subject variability on effect sizes, all participants significantly improved MVC strength in at least one joint during SCS ON compared to SCS OFF (**Extended Data Fig. 3a,b**). It is worth noting that the absence of a significant improvement in shoulder extension force may be attributable to our safety-driven protocol modification after the adverse event observed in SCS04 (**Extended Data Table 2**). Specifically, by avoiding stimulation through rostral contacts near the C3/C4 spinal levels, we may have limited the activation of motor pools of shoulder muscles (**Extended Data Fig. 1h,i**). Taken together, these results demonstrate that SCS can significantly increase strength regardless of impairment severity (**Supplementary Information Fig. 1)**.

### Impact of stimulation selectivity on strength outcomes

Selective activation of the appropriate spinal levels during each isolated movement was key to enable improvements in strength. For example, elbow extension torque was significantly increased in SCS04 when stimulation targeted the triceps brachii muscle using the 8R contact at the C7 spinal level (**Fig. 2d-f)**. In contrast, elbow extension was hindered when stimulation targeted the biceps brachii muscle using the 5R contact, which targeted more rostral (C5/C6) spinal levels (**Fig. 2d-f)**. Pooling data across all participants (**Fig. 2g,h**) allowed us to identify the spinal levels targeted by stimulation when participants showed improved strength at a given isolated movement. Specifically, using intraoperative electrophysiology data, we estimated which spinal levels were targeted by the contacts used to deliver SCS during the MVC task (**see Methods**). For instance, a significant increase in shoulder flexion MVC strength was associated with greater activation of the C4 spinal level, where motor pools innervating shoulder muscles are located (**Fig. 2g,h**). In contrast, improvement in grip MVC strength was associated with activation of the T1 level, where motor pools innervating hand muscles are located (**Fig. 2g,h**). These results support our hypothesis, indicating that the assistive effects of SCS depend on activating sensory fibers at spinal levels that innervate muscles involved in the intended movement. In other words, selective activation of appropriate spinal levels is critical for the efficacy of the assistive effects of SCS.

### Assistive effects of SCS on arm dexterity

We quantified the assistive effects of SCS on arm dexterity by measuring changes in the quality of arm kinematics during a planar reach-and-pull task when SCS was turned ON (**Fig. 3; Extended Data Figs. 4-6; Supplementary Information Fig. 2**). Specifically, the task was performed using an exoskeleton robot (KINARM, Kinarm) with anti-gravity arm support^31^ (**Fig. 3a; Supplementary Information Fig. 3; see Methods**). During the reach phase, participants were instructed to reach targets as straight as possible from a start position, focusing on movement control over speed (**Fig. 3b,c**). As exemplified in **Fig. 3a,b,e,f**, SCS resulted in significantly straighter and smoother reaching trajectories (**Fig. 3c**), indicating increased dexterous reaching movements. Importantly, all participants demonstrated significant improvement in arm reaching kinematics for at least one target with SCS ON (**Extended Data Fig. 5a-i**). During the pull phase, we observed similar improvements in arm kinematics under the SCS ON condition (**Extended Data Fig. 4a-d; Extended Data Fig. 6a-i**). Both reach and pull movements were significantly faster during the SCS ON condition (**Supplementary Information Fig. 4**). Notably, these assistive effects of SCS on arm kinematics were observed irrespective of whether or not participants showed increased strength in elbow flexion or extension during stimulation (**Extended Data Fig. 3**). This dissociation between the assistive effects on strength and arm dexterity suggests that SCS independently facilitates different neural drive modes during maximal versus submaximal muscle contractions^32^.

**Fig. 3.**
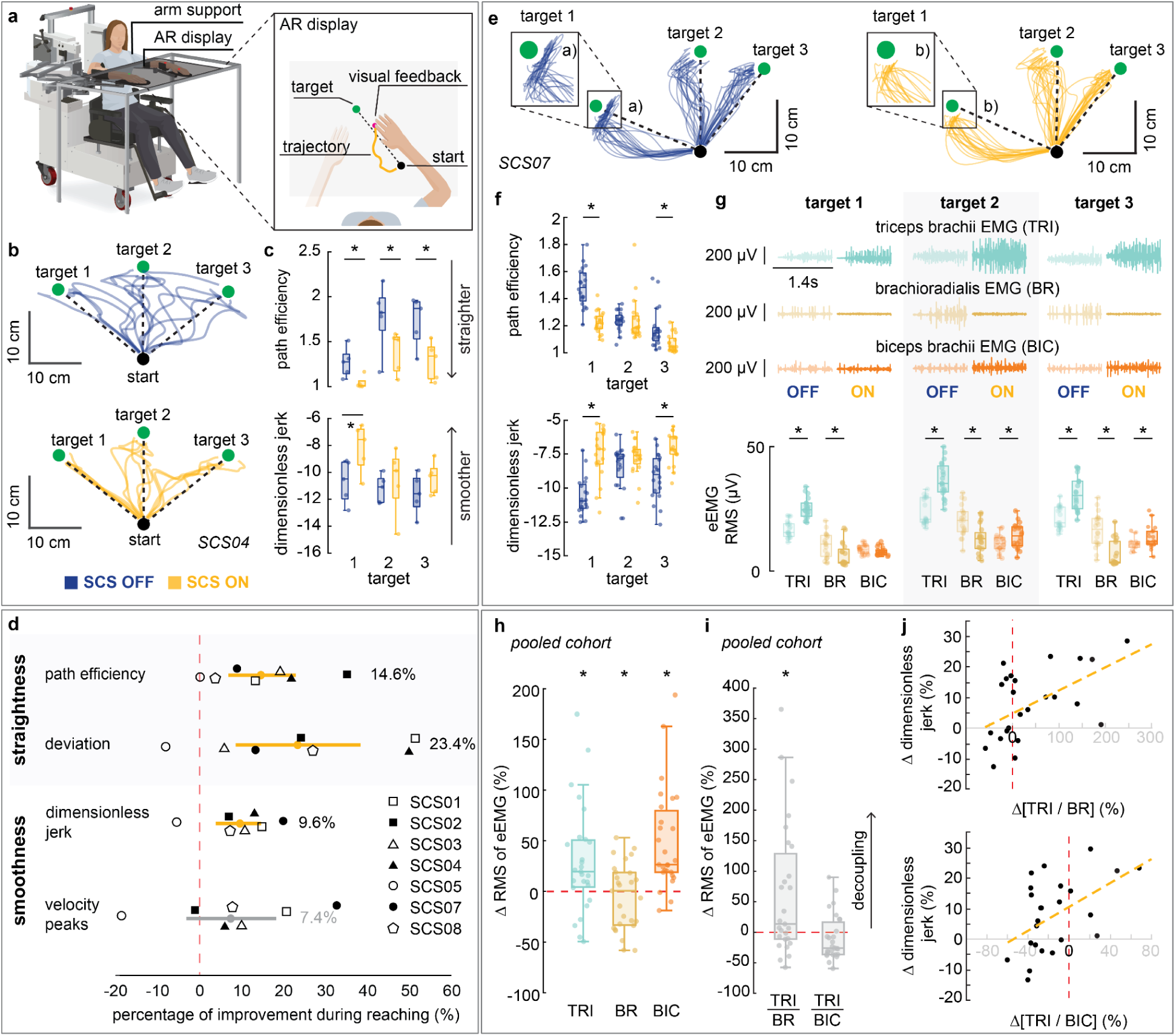
Assistive effects of spinal cord stimulation (SCS) on arm reach kinematics (dexterity). **a-c**. A planar reach-and-pull task was used to assess the assistive effects of SCS on upper-limb dexterity. **a.** During the reach phase, participants were instructed to reach an end target from an initial hand position. **b.** Example from participant SCS04 showing improved reach trajectories with SCS ON. **c**. Trajectories shown in panel b. significantly improved in straightness (path efficiency) and smoothness (log dimensionless jerk) metrics. **d**. Assistive effects of SCS on arm reach kinematics pooled across all participants. We observed significant improvements in all metrics except for the number of velocity peaks, suggesting overall improvements in dexterity. **e-g**. Trajectories, kinematics metrics, and electromyography (EMG) profiles for key muscles (triceps brachii (TRI), brachioradialis (BR), and biceps (BIC)) involved in the reach phase for SCS07. **e**. Trajectories were qualitatively straighter during the SCS ON condition compared to SCS OFF. In particular, trajectories near target 1 were noticeably straighter when stimulation was applied (a and b). **f**. Quantitative analysis confirms significant improvements in both straightness and smoothness during the SCS ON condition (assistive effects). **g**. EMG recordings showed increased activation of TRI and BIC, and reduced activity in BR during SCS ON (darker shades) compared to SCS OFF (lighter shades). Root-mean-square (RMS) of the EMG envelope (eEMG) was used to quantify muscle activity. **h-j**. Assistive effects of SCS on muscle synergies during reach phase. Asterisks in panels h and i indicate statistically significant differences between SCS ON and OFF conditions. **h.** Percentage change in RMS of eEMG signals from TRI, BR and BIC muscles between SCS ON and OFF conditions, pooled across all participants. Myoelectric activity of the TRI and BIC were amplified under SCS ON, while activity of the BR was suppressed. **i.** Agonist-antagonist activation ratios (TRI/BR and TRI/BIC) were used to quantify changes in muscle co-contractions between SCS ON and OFF conditions (SCS ON/ SCS OFF - 1). A positive ratio (> 0) indicates decoupling of the agonist-antagonist muscles. The agonist-antagonist ratio TRI/BR was significantly increased with SCS ON, whereas TRI/BIC was not statistically different across stimulation conditions. **j.** Correlation between the percentage change in agonist-antagonist muscle activation ratios (TRI/BIC and TRI/BR) and the corresponding percentage changes in log dimensionless jerk enabled by SCS during the reach phase. A significant positive linear correlation was observed for TRI/BR (r=0.530; p=0.004) and TRI/BC (r=0.502; p=0.006), indicating reduced co-contraction between the TRI and both antagonist muscles (BR and BIC) under SCS ON was associated with improved movement smoothness.

### Assistive effects of SCS on abnormal synergies

To analyze the of SCS on muscle synergies, we compared the myoelectric activity—quantified as the root mean square (RMS) of the EMG signal envelope (eEMG)—of elbow flexor and extensor muscles during the reach-and-pull task under SCS ON and OFF conditions. During the reach phase, myoelectric activity of the triceps brachii (TRI) and biceps brachii (BIC) muscles were amplified during SCS ON condition, whereas activity of the brachioradialis (BR) was suppressed (**Fig. 3g,h**). We then quantified agonist-antagonist co-contraction using activation ratios (TRI/BR and TB/BIC; see **Methods**). We found that TRI/BR, not TRI/BIC, was significantly increased during SCS ON, indicating a reduction of abnormal agonist-antagonist co-contraction (**Fig. 3i**). Notably, the increase in the TB/BR ratio was significantly correlated with improvements in arm kinematics (**Fig. 3j**; **Supplementary Information Table 6**). During the pull phase, a similar pattern of EMG activation was also observed, although activity of triceps brachii muscle tended to decrease during SCS ON condition (c.f., SCS02 as an outlier to this pattern in **Extended Data Fig. 4e,f**). In contrast to the reach phase, we found a significant decrease in TRI/BIC, not TRI/BR, which was significantly correlated with improvements in arm kinematics (**Extended Data Fig. 4g,h**). Together, these results suggest that SCS can modulate the activity of muscles acting at a single joint (elbow), thereby reducing abnormal muscle synergies and improving arm dexterity.

From complete plegia to isolated movements out-of-synergy, the impairment and recovery from abnormal joint synergies can be quantified through subcomponents of the FMA^33–35^. To assess the effects of SCS on abnormal synergies across joints, we analyzed scores of two FMA subcomponents: movement combining synergy (MCS) and movement out-of-synergy (MOS)^33–35^. Specifically, these subcomponents are aimed at evaluating the ability to individualize multi-joint movements outside of pathological coupling patterns observed after stroke^33–35^—for example, elbow extension during shoulder flexion. We found that assistive effects of SCS led to improvements in both MCS and MOS subscores (**Fig. 4a,b; Extended Data Fig. 7a-h**), suggesting reduction of abnormal synergies during SCS. Importantly, therapeutic effects observed over time in the absence of SCS showed complementary contributions in alleviating abnormal synergies, leading to marked facilitation of inter-joint movements as an effective improvement over the 4-week trial (**Fig. 4b**).

**Fig. 4.**
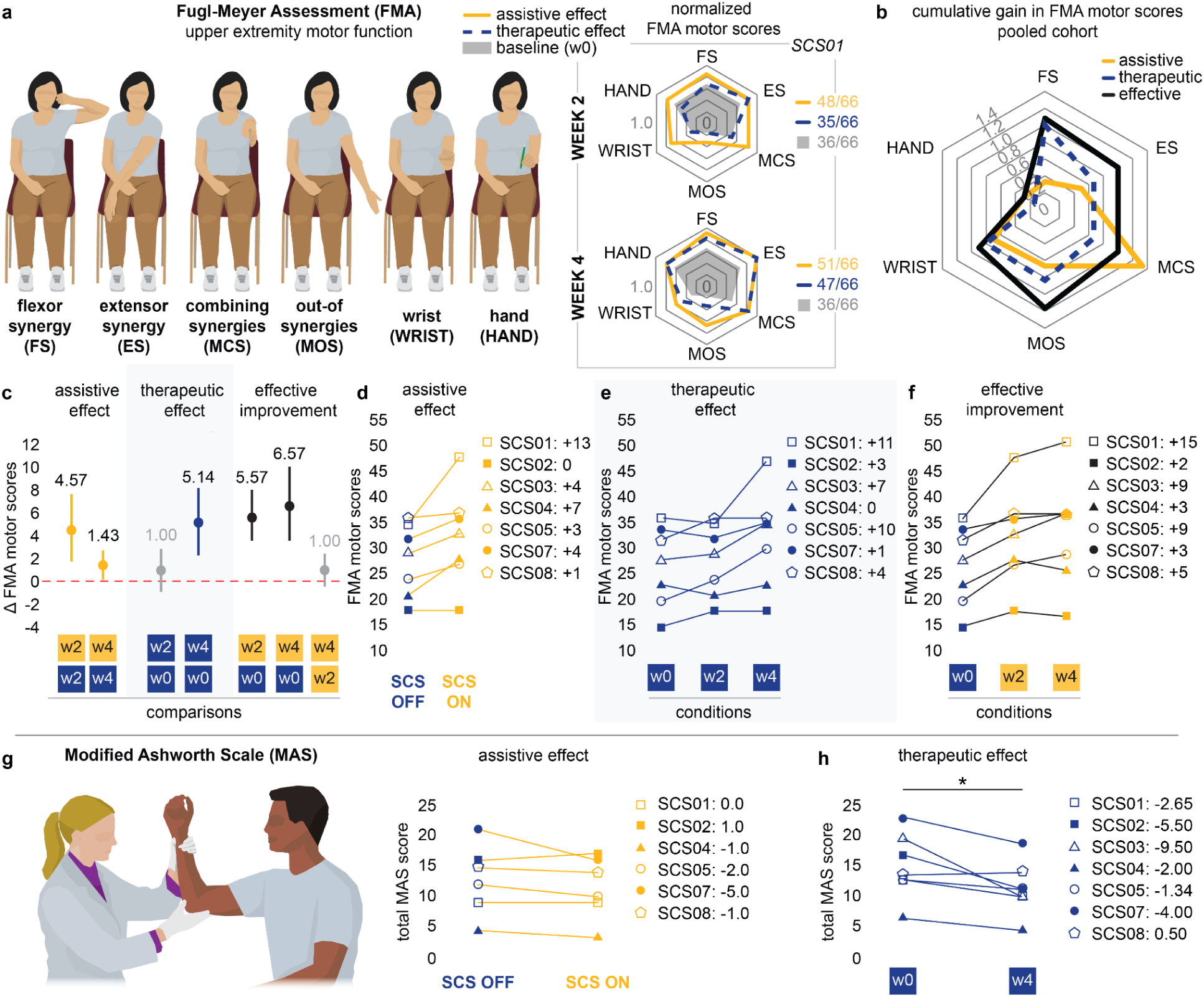
Clinical assessments quantifying the effects of spinal cord stimulation (SCS). **a-f.** The Fugl-Meyer Assessment (FMA) upper-extremity motor function was used to assess the effects of SCS. **a.** Web plots showing the normalized change in scores across subcomponents of FMA for SCS01. Baseline is indicated in shaded grey, assistive effects in yellow, and therapeutic effects in dashed blue. **b.** Cumulative change in FMA motor subcomponent scores across all participants. For each participant, improvements in subcomponent scores were normalized by the maximum score for that component (assistive effects, yellow; therapeutic effects, blue; effective improvement, black). **c.** Average change in the FMA motor scores throughout the trial across all participants. **d.** Assistive effects on FMA motor scores for each participant at week 2; **e.** Therapeutic effects on FMA motor scores measured during SCS OFF condition. **f.** Effective improvement of SCS on the FMA motor score. **g-h.** The Modified Ashworth Scale (MAS) was used for assessing spasticity. **g.** Assistive effects of SCS on the total MAS score. These assistive effects were assessed between weeks 2 and 4 (see Methods) and were not administered for participant SCS03. **h.** Significant therapeutic effects emerging over time on the total MAS score.

### SCS significantly improves total FMA motor scores

The Fugl-Meyer Assessment (FMA) is a reliable and validated clinical scale widely used to quantify impairment severity in participants with post-stroke hemiparesis and their recovery following therapeutic interventions^33,36^. We, therefore, employed the FMA to compare the effects of SCS (**Fig. 4a-f; Extended Data Fig. 7**) with previous studies^11,14,15^. Under the SCS OFF condition, we observed no significant change in FMA scores between baseline (pre-implant) and week 2, indicating weak therapeutic effects at the mid-study timepoint (**Fig. 4c**). In contrast, we measured a sizable increase in FMA scores when SCS was turned ON (mean +4.57 compared to SCS OFF, and +5.57 compared to baseline in **Fig. 4c**). These outcomes are consistent with the observed assistive effects of SCS on strength and arm dexterity, demonstrating that SCS can enable meaningful and immediate improvements, as quantified by a validated clinical scale. We then assessed changes in FMA scores at the end of week 4. At this end-study timepoint, FMA scores during the SCS OFF condition were significantly higher compared to baseline (mean +5.14 in **Fig. 4c**), indicating that participants retained improvements even in absence of stimulation, i.e., therapeutic effects. The effective improvement, reflecting the combined assistive and therapeutic effects, was even greater when comparing FMA scores during the SCS ON condition at week 4 and baseline (+6.57) (**Fig. 4c**). Surprisingly, the effective improvement at week 2 (+5.57) accounted for 85% of the maximum improvement observed at week 4 (+6.57), suggesting that just 2 weeks of SCS intervention were sufficient to produce most of the functional gains with stimulation ON (**Fig. 4c, Supplementary Information Fig. 5**).

### Effects of SCS on spasticity

Anecdotally, we noted that participants experienced an immediate reduction in muscle tone when SCS was turned ON. Participants often reported that these effects persisted after the experimental sessions, without stimulation. We then quantified the effects of SCS on spasticity using the Modified Ashworth Scale (MAS, **Fig. 4g-h**)^37,38^. During SCS ON, 5 out of 6 participants showed an immediate reduction in MAS score that exceeded the Minimal Clinically Important Difference (MCID) threshold (> 0.76) in at least one muscle group^39^ (**Fig. 4g, Extended Data Table 3**). Moreover, all participants experienced a clinically meaningful decrease in MAS scores from pre-implant (baseline) to week 4 (end of study) with SCS OFF in at least one muscle group (> 0.76; **Extended Data Table 3**)^39^. Importantly, these results indicate a strong and consistent effect of SCS in reduction in overall arm spasticity as a therapeutic effect of our intervention (**Fig. 4h**). Taken together, the effects of SCS on spasticity were surprisingly strong.

### Potential determinants of SCS responsiveness: an exploratory analysis

Given the observed variability in SCS effects across participants (e.g., **Fig. 4f**), we conducted an exploratory analysis to identify potential factors associated with the degree of effective improvement. We used total FMA motor scores to quantify changes in motor function in reference to other interventional approaches^14–16^. Particularly, the U.S.A. FDA recently approved VNS for motor rehabilitation in people with post-stroke hemiparesis based on reports showing a change of 5 points in the FMA motor scale after 6 weeks of intense rehabilitation^14^. Although the mean amount of physical activity in our protocol was about 30% of that in the VNS study (8.6 hrs versus 27 hrs), we adopted a 5-point FMA threshold to distinguish between high- and low-responders, consistent with the MCID for the scale^40^. We categorized participants with “high” (≥5) and “low” (<5) responsiveness to SCS based on their FMA scores at week 4 under the SCS ON condition relative to pre-implant (baseline), *i.e*., maximum effective improvement. The effective improvement in FMA motor scores was +9.5 points for high-responders and +2.7 points for low-responders. We then used their baseline FMA motor scores (**Fig. 5a**) and FMA sensory scores (**Fig. 5b**) to characterize their initial motor and sensory impairments, respectively. Our data suggested that baseline sensory function, rather than the initial motor impairment, may be a stronger determinant of responsiveness, i.e., significant effective improvement.

**Fig. 5.**
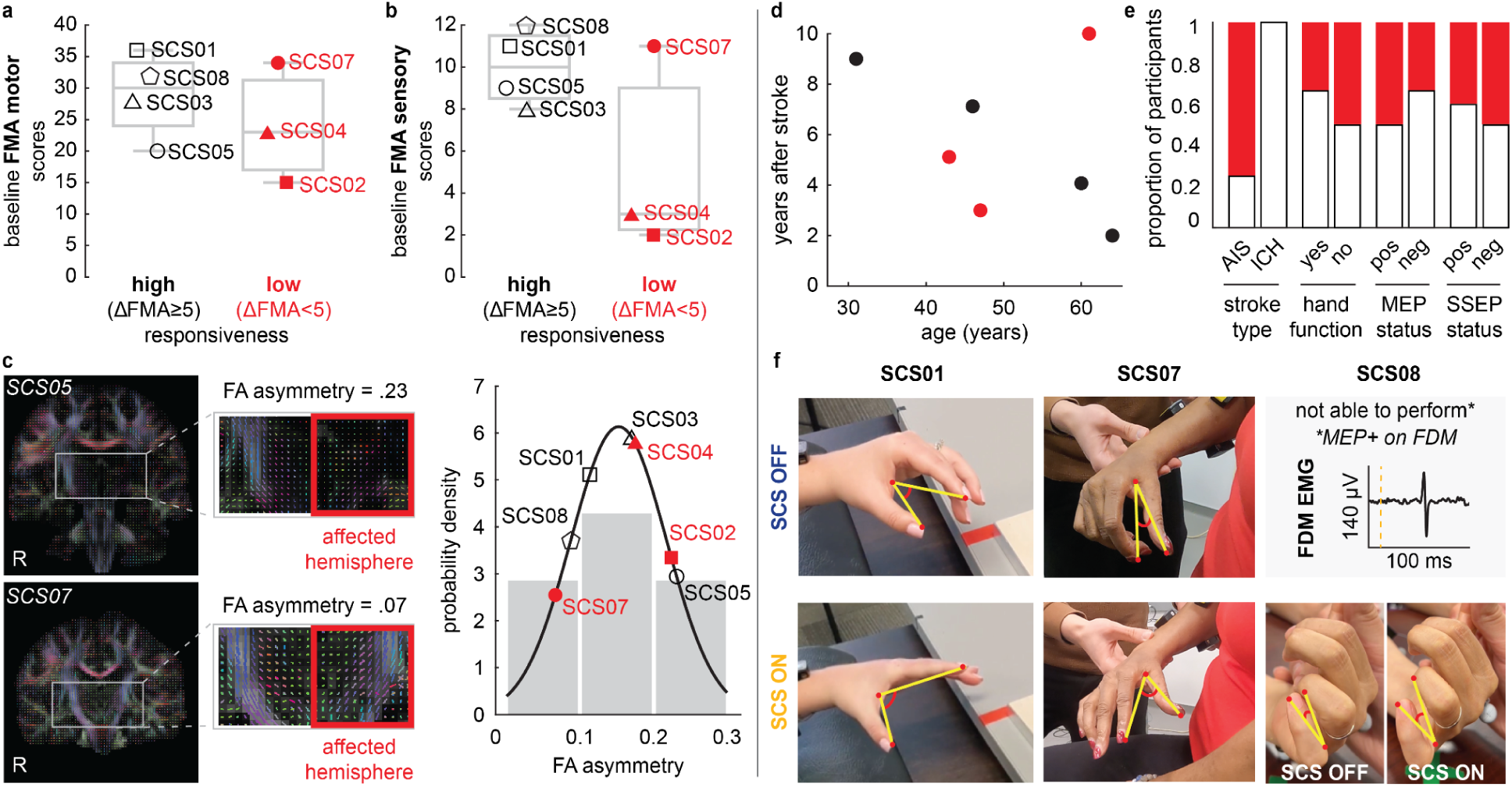
Exploration of potential determinants of spinal cord stimulation (SCS) responsiveness. **a-c.** Motor and sensory impairments as potential determinants of SCS responsiveness. **a.** Fugl-Meyer Assessment (FMA) scores for upper-extremity motor function and **b.** FMA scores for upper-extremity sensory function at baseline characterizing participants with “high” (ΔFMA≥5) and “low” (ΔFMA<5) responsiveness. ΔFMA was defined as the difference between FMA motor scores at baseline and at week 4 with SCS ON, i.e., effective improvement. Participants with “high” responsiveness are indicated in black, and those with “low” responsiveness in red. **c.** Diffusion-weighted MR imaging and tractography were used to assess corticospinal tract (CST) integrity via fractional anisotropy (FA) values. Representative tract profiles are shown for a participant with high FA asymmetry (SCS05) and one with low FA asymmetry (SCS07). The distribution of FA values across “high” and “low” responders is shown on the right. **d-f.** Other potential determinants of SCS responsiveness. **d.** Years after stroke and age of participants stratified by responder status. Black points indicate “high” responders, while red points indicate “low” responders. **e.** Proportion of high and low responders by stroke type (AIS versus ICH), hand opening ability assessed at pre-implant baseline, motor evoked potential (MEP) status and somatosensory evoked potential (SSEP) status. Abbreviations: AIS, acute ischemic stroke; ICH, intracerebral hemorrhage. **f.** Assistive effects of spinal cord stimulation (SCS) on hand and finger movements in participants with residual hand opening abilities at baseline or who were MEP+. Notably, SCS08 had no hand opening ability at baseline, but showed MEP+ responses in the flexor digiti minimi (FDM). She was able to move her 4th and 5th digits when SCS was ON.

We then examined the association between responsiveness to SCS and the residual anatomical integrity of the CST, quantified using fractional anisotropy (FA) of diffusion-weighted MR images (**see Methods**). Consistent with the weak association between baseline motor impairment and responsiveness in **Fig. 5a**, we found no clear relationship between CST integrity and effective improvement enabled by SCS (**Fig. 5c**). Finally, no strong associations were found for most of the other participant characteristics, except for stroke type (**Fig. 5d,e**). Notably, the weak association between MEP status and effective improvement on the FMA scale was also consistent with the above findings related to motor impairment and CST integrity. However, we observed that assistive effects on hand opening and finger individualization only occurred in the 3 out of 7 participants who had residual finger movement or were MEP+ (**Fig. 5f**). Consistent with **Fig. 4b**, hand and finger improvements were predominantly observed as assistive effects of SCS (c.f., HAND in **Fig. 4b**). These findings suggest that SCS can facilitate hand and finger control by facilitating residual direct monosynaptic CST connections^41^.

## DISCUSSION

Here, we report the final results of a pilot study aiming to assess the safety and preliminary efficacy of SCS in seven participants with profound post-stroke motor deficits (15 to 35 FMA score at enrollment). We demonstrated that cervical SCS can improve strength, dexterity, spasticity, and synergies with minimal physical exercise (only 8.6 hours) through a combination of assistive and therapeutic effects. Importantly, these improvements were consistent across all participants, regardless of the severity of their symptoms, or years after stroke. Moreover, we observed no serious adverse events, suggesting that SCS can safely deliver effective doses in people with stroke.

Neurostimulation approaches have been previously designed to catalyze therapeutic effects of motor rehabilitation—that is, stimulation is intended to act in combination with rehabilitation to promote changes in motor function over time^14–16^. In contrast, we demonstrated that SCS operates through a fundamentally different mechanism, immediately alleviating hemiparesis symptoms during stimulation, i.e., producing *assistive* effects. By facilitating cortico-spinal communication^42^, the combination of assistive and therapeutic effects resulted in significantly larger effect sizes across time. Notably, 85% of the maximum improvement in FMA scores observed at week 4 had already been achieved by week 2, when no significant therapeutic effects had been detected. These findings demonstrate that SCS alone, even without formal motor rehabilitation, can achieve effect sizes comparable to other effective neurostimulation approaches^15^, including FDA-approved VNS^14^. Importantly, the total amount of active movement performed by our participants was, on average, at least 3 times lower than other neurostimulation trials^14,15^ (8.6 hours) and similar to the standard-of-care in the outpatient setting (7 hours)^13^. This emphasizes the rapid and substantial benefits of assistive effects of SCS, and suggests that combining SCS with a high-dose motor rehabilitation program could lead to even greater improvements^12,43,44^. It is also worth noting that the cohort included in this clinical trial would be ineligible in most other interventional studies. For example, an appreciable proportion of the participants in our study were MEP-negative^15^ across arm muscles, did not have residual finger function^12,14^, and had low baseline FMA sensory scores (<6)^14^.

Although facilitation of hand and finger movements strongly depended on residual direct monosynaptic CST connections to finger muscles, this condition was not a prerequisite for motor improvements in strength, dexterity, abnormal synergies and spasticity. Indeed, using the FMA scale as an index quantifying motor improvements in the arm and hand, our data suggested that spared sensory function—rather than motor function—was a potential determinant of responsiveness to SCS. Consistent with these results, improvements in arm strength and dexterity did not appear to depend on baseline motor impairments either. For instance, although SCS02 did not regain finger function, she showed substantial improvements in strength, arm dexterity, and spasticity. Notably, SCS02 would have been ineligible in all aforementioned trials^12,14,15^. Importantly, while most therapies have been tested in individuals with ischemic stroke^14,15,45^, we found that SCS is also effective for those with hemorrhagic stroke. These results demonstrate that SCS is agnostic to the cause of CST damage, suggesting its rehabilitative potential in other neurological disorders with pathophysiological similarities to hemorrhagic stroke, such as traumatic brain injury^46^.

Furthermore, we observed consistent improvements in spasticity over time, with all participants showing significant reductions in MAS scores. Participants often self-reported these changes. While previous studies have suggested that SCS may reduce spasticity in other neurological conditions^47,48^, we were nonetheless surprised to observe these effects emerge over time in the absence of stimulation. Although SCS is generally thought to excite neural circuits, this inhibitory effect on spasticity may reflect homeostatic adaptations after prolonged use of stimulation^27^. Importantly, these strong outcomes support the potential of SCS as an alternative to standard, and often costly, spasticity treatments^49^, such as repeated botulinum toxin injections^50^, pharmacological treatments^51^, and implanted intrathecal baclofen pumps^52^. Additionally, unlike SCS, these spasticity treatments can also impair motor control because they inhibit muscle activity^53^.

In this study, we also analyzed the impact of stimulation parameters on effect size. We demonstrated that selective stimulation of appropriate spinal levels is critical for achieving meaningful assistive effects, which is an important consideration for testing alternative non-invasive stimulation approaches^54,55^. For example, selective stimulation of caudal dorsal roots (C8/T1) was critical to facilitate hand function. In this regard, here we present a reproducible method to rapidly identify and configure selective stimulation contacts, making this feasible for clinical implementation. Using this approach, intraoperative neuromonitoring teams can assist surgeons in optimizing lead placement and identifying selective contacts, while physical therapists can efficiently fine-tune SCS parameters by evaluating immediate changes in motor function.

This study is limited by two factors. First, the small sample size restricts the validity of our quantitative estimation of the effect size and limits the ability to explore determinants of responsiveness. However, the heterogeneity of our cohort points to the versatility of SCS in a broad stroke population. Second, the absence of a control group performing only motor assessments restricts the interpretation of our results with regard to therapeutic effects emerging over time in the absence of stimulation. However, this limitation does not affect the bulk of our analysis, as most efficacy outcomes focused on the assistive effects of SCS, with each participant serving as their own control under SCS OFF condition. Even though we did not execute formal control for the therapeutic effects, the results of the VNS trial allow us to speculate on this point. Indeed, in the VNS study, control participants underwent a formal rehabilitation program of 27 hours that led to average +2.4 points on the FMA motor scale^14^. Using this value as a statistically powered reference, we observed a contrasting result in our study: participants showed a mean improvement of +5.14 FMA motor points under SCS OFF after 4 weeks, despite performing minimal active movement (a total of 8.6 hours)—consistent with the current standard of care^13^. Therefore, our results suggest that the observed therapeutic effects are largely attributable to the SCS intervention. Future clinical studies should address these limitations by performing direct comparisons with controls in larger populations.

In conclusion, our results suggest that SCS could be an effective, reproducible, scalable and clinically viable neuroprosthetic technology for treating movement dysfunctions and spasticity in post-stroke hemiparesis.

## METHODS

### Study design and cohort

#### Trial and participant information

All experimental protocols were approved by the University of Pittsburgh Institutional Review Board (IRB) (protocol STUDY19090210) under an abbreviated investigational device exemption. The study protocol is published on ClinicalTrials.gov (NCT04512690). Eight participants were enrolled in the study, however only seven completed the study. Specifically, SCS06 was withdrawn from the trial after being diagnosed with arrhythmia during preoperative assessments. All participants provided informed written consent according to the procedure approved by the IRB of the University of Pittsburgh and participants were compensated for each day of the trial and for travel and lodging during the study period.

#### Inclusion criteria

Individuals between 21 and 70 years of age who had suffered from an ischemic or hemorrhagic stroke more than six months prior to the start of the study were eligible for participation. All individuals had hemiparesis affecting their upper limb with baseline Fugl-Meyer Assessment (FMA) scores for the upper-extremity motor function between 7 and 40. Before the study, participants were screened with a rigorous medical evaluation. Candidates with severe comorbidities, previously implanted medical devices, claustrophobia, pregnant or breastfeeding were excluded from the study. Individuals were not receiving any anticoagulant, anti-spasticity, or anti-epileptic medications for the duration of the study period.

#### Study design and data reported

The goal of this pilot clinical trial is to evaluate the safety and efficacy of SCS for improving motor function in individuals with chronic post-stroke upper-limb hemiparesis. The study is designed as a single-group, open-label, prospective study. Given the pilot nature of the study, to minimize safety risks, SCS leads are implanted for a maximum period of 29 days, after which the electrodes are explanted. We designed our primary and secondary outcomes to primarily assess safety and obtain preliminary clinical and scientific evidence of both the assistive and long-term effects of SCS.

After screening and pre-study baselines, participants are implanted with clinical SCS leads. Starting from day 4 post-implant, participants undergo scientific sessions five times per week, 4 hrs per day, for a total of 19 sessions until explantation day. Tasks and measurements during the first week are focused on identifying optimal stimulation configurations that are then maintained for the remaining sessions. A detailed description of primary and secondary outcomes can be found in **Supplementary Information Table 1** and on ClinicalTrials.gov (NCT04512690).

In summary, the primary outcome of this study is safety, which was assessed by systematically reporting serious adverse events. Specifically, we consider the trial to be successful if there are no serious adverse events related to the use of the stimulation. We asked participants to report and rate, if present, any pain or discomfort that arises from the stimulation with the goal of understanding if intensities required for motor effects are pain- and discomfort-free. The secondary outcome of this study is the efficacy of SCS for improving motor function. We assessed the efficacy of SCS in terms of (*i*) assistive effects, defined as the difference between SCS ON and SCS OFF conditions at any timepoint; (*ii*) therapeutic effects, defined as the difference between timepoints for SCS OFF condition; and (*iii*) effective improvement, defined as the difference between pre-implant baseline and SCS ON condition at any timepoint. Specifically, we quantified the assistive effects of SCS on strength, by measuring maximum voluntary contraction (MVC) forces during isometric movements; dexterity, by analyzing arm kinematics during a planar reach-and-pull task; spasticity, using the Modified Ashworth Scale (MAS) across multiple upper-limb joints; and abnormal synergies, by comparing electromyography (EMG) signals of elbow antagonist and agonist muscles during a reach-and-pull task and subcomponents of the FMA for upper limb motor function. We also quantified the therapeutic effects emerging over time on spasticity, using the MAS; and abnormal synergies, by comparing subcomponents of the FMA. Moreover, we assessed the effective improvement enabled by SCS using total FMA motor scores, thereby enabling comparison of our effect sizes with those reported in the literature^11,14,15^. Finally, we also performed a battery of imaging and electrophysiology tests to assess motor and sensory impairments at baseline. Below we detail the methods for each of the measurements reported in this trial.

#### Participant information

SCS01 is a female who had a right hemorrhagic stroke secondary to a thalamic/midbrain cavernous malformation nine years before participating in the study. At the time of the study, she had left-sided spastic hemiparesis. Her spasticity was previously managed with botulinum neurotoxin injections to the biceps, brachioradialis, and pronator teres. These injections were suspended six months prior to and throughout the study. She had also undergone flexor tendon lengthening surgery and a C5–C6 anterior cervical discectomy and fusion in the interim. SCS01 was able to partially open her hand and perform some individualized movements with her 1st and 2nd digit.

SCS02 is a female who had a right ischemic MCA stroke secondary to a right carotid dissection resulting in a large MCA territory infarct 3 years before participating in the study. Her post-stroke residual at the time of participation was a left-sided spastic hemiparesis complicated by a left wrist flexion contracture despite treatment with splinting. SCS02 was unable to open her hand or perform individualized finger movements.

SCS03 is a male who had a hemorrhagic stroke in the right MCA territory, followed by craniotomy and hematoma evacuation seven years prior to the study. At the time of study participation, he had left-sided spastic hemiparesis. He was not taking medications for spasticity but used a home shockwave device for spasticity management and actively attended outpatient physical therapy three times per week. SCS03 was unable to open his hand or perform individualized finger movements.

SCS04 is a male who had an ischemic stroke in the right MCA territory five years prior to study participation. He presented with left-sided hemiparesis and underwent thrombolysis. He completed both inpatient and outpatient rehabilitation and was not being managed for his spasticity at the time of the study. SCS04 was unable to open his hand or perform individualized finger movements.

SCS05 is a male who had a left basal ganglia intraparenchymal hemorrhage two years prior to study participation, resulting in right-sided hemiparesis and transient speech difficulties. MRI confirmed a basal ganglia hemorrhage without underlying vascular abnormalities, consistent with a likely hypertensive etiology. His post-stroke rehabilitation included physical, occupational, and speech therapy with limited functional improvement. He received botulinum neurotoxin injections for spasticity, which were discontinued 10 weeks prior to and throughout the duration of the study. SCS05 was able to partially open his hand.

SCS07 is a female who sustained a left hemispheric lacunar infarct ten years prior to study participation, with a past medical history of hypertension and diabetes mellitus. Her post-stroke deficits included weakness and spasticity in her right upper limb. She actively participated in physical and occupational therapy prior to the study. She also had a joint contracture in the shoulder joint. She weaned off baclofen prior to enrolling in the study. SCS07 was able to partially open her hand and perform some individualized movements with her 1st and 2nd digit.

SCS08 is a female who had a right-sided ischemic stroke involving the corona radiata four years prior to the study. She has a notable past medical history of hypertension. Her post-stroke deficits include persistent weakness and spasticity in the left upper and lower extremities, with preserved sensation in the affected arm. She underwent inpatient rehabilitation following her stroke and was followed by 6 months of at-home physical and occupational therapy. She was managed with continual botulinum neurotoxin injections for left hand and thumb tightness, but these were held for four months prior to and during the trial. At the pre-study baseline, SCS08 was unable to open her hand or perform any individualized finger movements.

### Safety

We systematically documented all adverse events using a log form provided by the Institutional Review Board (IRB) of the *University of Pittsburgh* and developed by the *Education and Compliance Office for Human Subject Research*. All adverse events were reported to the Data Safety and Monitoring Board (DSMB) of the University of Pittsburgh, as well as to the IRB, to determine whether or not they would be related to the clinical trial. Throughout this clinical trial, we strictly followed all IRB directives for participant safety. For instance, following a study-related adverse event with SCS04, we rigorously adhered to IRB directives by reporting the event to the FDA and revising our stimulation protocol to ensure participant safety.

### Intraoperative workflow/placement of leads

For specific information regarding the surgical procedure, see **Methods in Supplementary Information**.

#### Intraoperative recruitment curves

To confirm placement of each electrode lead and ensure activation of proximal arm muscles to distal hand muscles, we measured recruitment curves during the surgical procedure. Specifically, stimulation was delivered in a monopolar configuration using an intraoperative neuromonitoring system (Xltek Protektor, Natus Medical). A subdermal return electrode was placed at the deltoid or upper back contralateral to the affected arm of the participant. For each SCS contact, stimulation pulses were delivered at 1-2 Hz with amplitude ramped up from 0 to 5 mA. Compound muscle action potentials (CMAPs) were recorded using subdermal needle electrodes bilaterally in both arms. EMG was often recorded from trapezius, anterior deltoid, medial deltoid, posterior deltoid, biceps, triceps, pronator teres, wrist flexors, wrist extensors, abductor pollicis, abductor digiti minimi. We recorded contralateral myoelectric activity to ensure that SCS did not induce cross-over effects to the contralateral arm.

### Electrophysiology assessments

#### Motor evoked potential (MEP) status

Motor evoked potential (MEP) status was used to assess the integrity of the corticospinal tract (CST) after stroke^56,57^. To determine the MEP status, transcranial magnetic stimulation (TMS) with a Magstim 200^2^ and 70 mm D figure-eight coil was applied at a 3 x 3 cm *hotspots* grid centered over the hand knob area of the primary motor cortex in the affected hemisphere. Participants were classified as MEP-positive (MEP+) if peak-to-peak MEP amplitudes were ≥50µV in either proximal arm muscles (i.e., deltoid, biceps, triceps) and/or distal hand muscles (i.e., abductor pollicis brevis, abductor digiti minimi, flexor digiti minimi) contralateral to the stimulated cortex. Participants were classified as MEP-negative (MEP-) if no MEPs could be evoked.

#### Somatosensory evoked potential (SSEP) status

Somatosensory evoked potential (SSEP) status was used to assess the integrity of ascending sensory pathways^56,58^. Specifically, upper extremity SSEPs were acquired intraoperatively during lead placement through subdermal needle stimulation of the ulnar nerve in the affected arm and subsequently recording from the contralateral cortical hemisphere using scalp electrodes placed according to the standard international 10-20 system^59^. The cortical responses were specifically recorded from Fz-P4 and Fz-P3 electrode montages. Stimulation frequency was 3.11 Hz with a single pulse duration of 0.3 msec. Bandpass filters were set at 10-250 Hz with a maximum range window of +/- 2.5 mV. Participants were classified as SSEP-positive (SSEP+) if the peak-to-peak SSEP amplitudes were ≥ 5µV, and as SSEP-negative (SSEP-) otherwise.

### Hand opening ability

Hand opening ability was assessed at baseline, prior to SCS leads implantation, using the Fugl-Meyer Assessment (FMA) for upper-extremity motor function (see below). Participants who scored > 0 points on the finger mass extension subitem were considered to have hand opening ability—that is, partial or full voluntary extension of the stroke-affected hand.

### Imaging

#### Intraoperative fluoroscopy

Intraoperative anterior-posterior and lateral fluoroscopic images (C-arm) were used to guide accurate percutaneous placement of the SCS leads. These fluoroscopic images were used to estimate the relative position of the electrode leads along the cervical spinal cord, and to develop a spinal segment map.

#### X-Ray

Anterior-posterior and lateral X-Rays were taken 1-2 weeks post-implant to evaluate for lead stability. We used these X-rays to estimate the relative position of the electrode leads along the cervical spine in **Extended Data Fig. 1**.

#### Structural Magnetic Resonance Imaging (MRI)

Magnetic resonance imaging (MRI) was acquired using a 3T Prisma system (Siemens) using a 64-channel head and neck coil. A T1-weighted structural image was captured using a magnetization-prepared rapid gradient echo sequence (repetition time = 2,300 ms; echo time = 2.9 ms; field of view = 256×256 mm^2^; 192 slices, slice thickness = 1.0 mm, in-plane resolution = 1.0×1.0 mm^2^). Additionally, a T1-weighted FLAIR sequence was captured (repetition time = 5,000 ms; echo time = 390 ms; field of view = 256×256 mm^2^; 192 slices, slice thickness = 1.0 mm, in-plane resolution = 1.0×1.0 mm^2^) and was ultimately used to depict the lesions in **Fig. 1d**. Lesion segmentation was performed manually for each slice of the sequence using the MRIcron image viewer (NITRC); the resulting region of interest was smoothed on all planes using a Gaussian smoothing kernel with full-width at a half-maximum of 2 mm. MRIcroGL (NITRC) was used to visualize and export the resulting segmented overlays.

#### Diffusion Tensor Imaging (DTI) for tractography

To assess the anatomical integrity of the corticospinal tract (CST), we estimated the asymmetry of fractional anisotropy (FA) between the lesioned and non-lesioned hemispheres using diffusion tensor imaging (DTI). A 3T MRI scanner was configured to use a diffusion spectrum imaging scheme to capture a total of 257 diffusion samples. The maximum b-value used was 4,000 s mm^2^ and the in-plane resolution and slice thickness were 2 mm. A diffusion sampling length ratio of 1.25 was used. The output resolution in diffeomorphic reconstruction was 2 mm isotropic. The restricted diffusion was quantified using restricted diffusion imaging^60^. The tensor metrics were calculated and a deterministic fiber tracking algorithm was used to reconstruct the CST fibers^61^. A region-of-interest (ROI) was placed in the midbrain, and a seed region was defined in Brodmann area 4 (BA4). For fiber tracking, we used an anisotropy threshold of 0.01, an angular threshold of 65 degrees. The step size was set to voxel spacing. Tracks with lengths shorter than 30 mm or longer than 300 mm were discarded. A total of 10,000 seeds were calculated for both hemispheres. Tracts were manually pruned after the execution of the algorithm. We then calculated the mean FA values for the left and right CST. The asymmetry between FA values calculated for the lesioned and non-lesioned hemispheres was computed using Stinear’s formula:

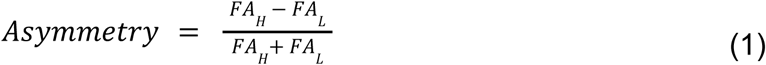

where *FA_L_* is the mean FA value of the CST in the lesioned hemisphere and *FA_H_* is the mean FA value of CST in the intact hemisphere.

### Calibration of stimulation parameters

#### Recruitment curves

To evaluate the selectivity of SCS in recruiting individual muscles, we estimated recruitment curves by measuring the peak-to-peak amplitude of compound muscle action potentials (CMAP) as a function of stimulation amplitude. We used a wireless EMG system (Delsys, Trigno) to record the CMAPs from multiple upper-limb muscles, such as trapezius, anterior deltoids, biceps brachii, triceps brachii, brachioradialis, abductor pollicis brevis, and adductor digiti minimi. Stimulation was delivered to one contact at a time at 1-2 Hz, with amplitudes gradually increasing in steps from 0.2 mA up to 8 mA, or until the participant reported discomfort. Monopolar stimulation was applied using a return surface electrode placed over the iliac crest contralateral to the affected arm. We measured the peak-to-peak amplitude of the CMAPs elicited at each stimulation amplitude. For each muscle, we normalized the amplitude of CMAPs to the maximum amplitude obtained across each measured trial. We repeated this process for each contact on both leads, thereby obtaining recruitment curves that described a relationship between SCS amplitude and muscle recruitment (e.g., **Fig. 2a; Extended Fig. 2**).

#### Systematic procedure for SCS calibration

Following a systematic procedure, we calibrated the stimulation parameters of a set of contacts to facilitate voluntary movements across multiple upper-limb joints. First, we used muscle recruitment curves to select contacts that selectively activated different muscle groups (e.g., elbow flexors vs. extensors). For each selected contact individually, we tuned the stimulation amplitude and frequency while participants performed movements involving the joint(s) targeted by stimulation (e.g., elbow flexion vs. extension). For example, when testing caudal contacts targeting the C8/T1 spinal levels, we assessed hand function and grip strength. Stimulation amplitude was increased in a range between 0.2 mA and 8 mA, and frequency between 40 Hz and 100 Hz. Pulse-width was typically set to 200 µs. Frequency was only increased when increasing stimulation amplitude failed to produce satisfactory outcomes. After individually calibrating a set of contacts (usually 2 or 3), we combined them with a personalized configuration for each participant. Stimulation amplitudes were re-adjusted when combining multiple contacts to minimize interference between their effects. Less frequently, we also adjusted the stimulation frequency across contacts.

### Maximum Voluntary Contractions (MVC) during isolated movements

Maximum Voluntary Contraction (MVC) force produced during isometric movements of the shoulder and elbow joints was assessed using a robotic torque dynamometer (HUMAC NORM, CSMi). During testing, the robot’s manipulandum was locked at a predetermined angle, and participants were instructed to exert their maximal voluntary force in either flexion or extension during 5 seconds. The joint under evaluation, shoulder or elbow, was aligned with the rotation axis of the manipulandum. To ensure isolation of single-joint motion, participants were stabilized using shoulder harnesses and joint-specific straps and braces according to HUMAC NORM’s recommended configurations. For example, during elbow strength testing, the upper arm and elbow were firmly supported against the chair back, and the forearm was held at a 90° angle to prevent movement. Each movement was assessed over 5 to 6 repetitions, with 10-15 second rest between repetitions. A 2 minute-break was often provided after the third repetition. The maximum torque value from each repetition was considered for analysis.

Grip strength was similarly quantified using a hydraulic hand dynamometer (Fabrication Enterprises, no. 12-0021). Participants were instructed to hold the device in a comfortable position and exert maximum grasping force for 5 seconds. The highest value from six repetitions was recorded and used for statistical analysis. Assistive effects of SCS on strength were assessed by comparing MVC force under SCS ON and OFF conditions within the same experimental session.

MVC testing was also performed on the unimpaired limb using the same procedures and joint configurations to evaluate for the degree of strength impairment across participants.

### Selectivity index

We assessed the association between the motor neuron pool targeted by SCS during the MVC task and the corresponding improvements in upper limb strength. First, we mapped the spinal levels targeted by the SCS parameters used during the MVC task described above. We then estimated a selectivity index for each spinal level, quantifying the degree to which activation of that level was associated with increased MVC strength across each isolated movement.

#### Spinal segment maps

For each participant, we mapped the location of upper limb motor neuron pools across the cervical spinal levels, producing individual spinal segment maps. We used intraoperative fluoroscopic X-Rays to identify the contacts positioned closest to the dorsal root fibers at each spinal level from C4 to T1. For each of these contacts, we analyzed intraoperative recruitment curves (see Methods for recruitment curves above) to identify the muscles activated at motor threshold—defined as the lowest stimulation amplitude delivered by each contact that evoked CMAPs with peak-to-peak amplitudes ≥ 50 µV. In some cases, multiple muscles were simultaneously activated at motor threshold amplitude. This process was repeated across all contacts. We then mapped the most selectively activated muscles, i.e. those activated at motor threshold, across spinal segments **(Extended Data Fig. 1a-g)**. Finally, we generated cohort-level spinal maps by overlapping the activated spinal segments for each muscle across all participants (**Fig. 2g**).

#### Selectivity index

A spinal selectivity index was developed to identify the spinal levels that contributed to increases in MVC force across all participants **(Fig. 2g,h)**. We used X-ray images acquired at week 2 post-implant to obtain an accurate estimation of the relative position of the leads along the spinal cord during the MVC task. We then identified the spinal levels with dorsal root fibers in closest proximity to the contacts used during the MVC task, i.e., spinal levels most targeted by the contacts used. For each contact, we computed a selectivity index, defined as the ratio between the stimulation amplitude used during the MVC task and the motor threshold amplitude estimated for the targeted spinal levels (see Methods for spinal segment maps above). The selectivity index was computed for the stimulation configurations used during the MVC task of each isolated movement (shoulder flexion, shoulder extension, elbow flexion, elbow extension and grip). For each spinal level and movement, selectivity indices were averaged across all participants who showed a significant increase in MVC force (**Extended Data Fig. 3**). Finally, the selectivity indexes for each isolated movement were normalized by the sum across spinal levels to generate the heatmap in **Fig. 2h**.

### Arm kinematics during reach-and-pull task

To assess the assistive effects of SCS on dexterity, we quantified arm kinematics during a planar reach-and-pull task. Specifically, we used a robotic augmented reality exoskeleton system (KINARM, Kinarm). Participants were secured in a modified wheelchair and their arms were suspended in the exoskeleton to remove the effects of gravity^62^. The platform displayed virtual targets onto a dichroic augmented reality display in front of the individual that allowed them to visualize their hand position relative to the virtual graphics.

The assessment task consisted of two phases: reach and pull. At the beginning of each trial, the participant placed their hand on the starting position. A target then was presented, followed by an audio cue after a randomized 100–700-ms delay, indicating the start of the reach phase. During the reach phase, participants were instructed to reach as straight as possible towards the target, focusing on movement control over speed. A target was considered successfully reached if the participant held the hand position within a 0.5-cm radius of the target center for 500 ms. Upon successful reach, an audio cue indicated the end of the reach phase. If the participant was unable to reach the target, the assessment continued with the next trial. An audio cue was played indicating the start of the pull phase. During the pull phase, participants were instructed to return their hand to the starting position without any specific instructions about the movement performance. Each target was presented 5 to 10 times in random order. For each participant, appropriate targets were selected based on their individual range of motion.

During reach and pull phases, hand trajectories and velocity were analyzed using metrics that characterize movement straightness (path efficiency and deviation error) and smoothness (log dimensionless jerk and velocity peaks)^63^, respectively **(Supplementary Information Fig. 3)**. Trajectories were low-pass filtered with a zero-lag, sixth-order Butterworth filter with a 10 Hz cutoff. To isolate arm kinematics during movement, we analyzed trajectory segments between 1.5 cm from the start (reach), or target (pull), position and 2.0 cm from the target (reach), or start (pull), positions. The following metrics used to quantify arm kinematics during planar reach and pull:

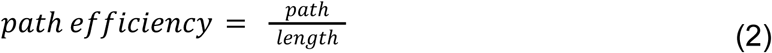

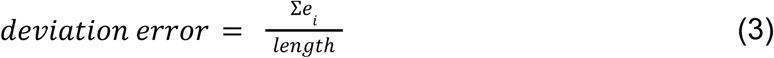

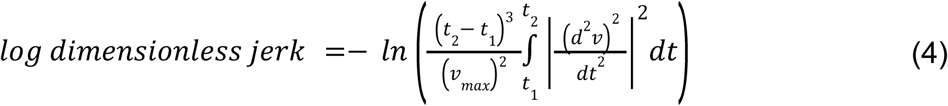

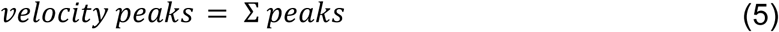

where *path* is the trajectory length (2), and *length* is the minimum Euclidean distance between the initial and end positions (2 and 3); *e*_*i*_ is the minimum distance between the ith sample point of the trajectory and the line defined by the center of the start and target positions (3); *v* is the velocity of the hand trajectory as a function of time, defined between *t*_1_ and *t*_2_, and *v*_*max*_ is the maximum hand velocity (4); and *peaks* is the number of velocity peaks with a minimum peak height of 0. 05 * *v*_*max*_ (5). Overall, the quantitative measures of straightness were used to assess the geometric deviation of the trajectories from a straight line, whereas the smoothness metrics were used to capture changes in hand acceleration and deceleration during reaching, independent of trajectory geometry. Additionally, the durations of the reach and pull phases were calculated. Assistive effects of SCS on dexterity were assessed by comparing arm kinematics during the reach and pull phases, characterized by four metrics, under SCS ON and OFF conditions within the same experimental session. The same SCS parameters were used for reach and pull phases (**Supplementary Information Fig. 2**). The task was repeated 1 to 4 times per stimulation condition, with a rest break (2-5 minutes) given between trials. Stimulation conditions were pseudorandomized. Targets with at least five successful reach trajectories were included in the analysis.

### Clinical assessments

#### Fugl-Meyer Assessment (FMA) for upper-limb motor function

The FMA is a reliable and validated clinical scale widely used to quantify impairment severity in participants with post-stroke hemiparesis and their recovery following therapeutic interventions^33,36^. We, therefore, employed the FMA to enable comparison of our effect sizes with those reported in previous studies^11,14,15^. The FMA for upper-extremity motor function consists of 33 items, each scored on a scale from 0 to 2, where higher scores indicate better motor execution (maximum score is 66). A trained occupational therapist (A.B.) conducted all assessments. Video recordings of the assessment were taken and reviewed to confirm results. The FMA was administered at baseline (pre-implant), week 2 post-implant, week 4 post-implant and follow-up (a month post-explant) timepoints. At week 2 and 4, the FMA was conducted under SCS ON and OFF conditions within the same experimental session. The order of stimulation conditions was pseudorandomized, with a 5-10 minute rest break provided between them. The evaluator was blinded to the stimulation condition. A minimal clinically important difference (MCID) of 5 points was used to interpret meaningful change^14,40^. Assistive effects of SCS were assessed by comparing total FMA motor scores between SCS ON and OFF conditions within the same session (week 2 and 4). Therapeutic effects were assessed by comparing FMA scores under SCS OFF across timepoints (baseline, week 2, week 4 and follow-up). Effective improvements were assessed as the difference in FMA scores between SCS OFF at pre-implant and SCS ON at week 2 and 4.

#### Fugl-Meyer Assessment (FMA) motor subcomponents

For each participant, we also analyzed the scores of 6 subcomponents of the FMA motor across stimulation conditions and timepoints: Flexor Synergy (FS), Extensor Synergy (ES), Movements Combining Synergies (MCS), Movements Out-of-Synergy (MOS), Wrist (WRIST) and Hand (HAND). For visualization in web plots (**Fig. 4a** and **Extended Data Fig 7**), the subscores for each participant were normalized to the maximum score of the corresponding subcomponent.

As a complementary analysis to assess the effects of SCS on the FMA motor subcomponents in the pooled cohort, we calculated the difference between the subscores across stimulation conditions and timepoints (**Fig. 4b**). Specifically, assistive effects of SCS were assessed by comparing FMA subscores between SCS ON and OFF at week 2, when assistive effects on total FMA motor scores were on average maximal and confounding influence from therapeutic effects was minimal. Therapeutic effects were assessed by comparing FMA subscores under SCS OFF between baseline and week 4, when therapeutic effects on total FMA motor scores were maximal. Effective improvements were assessed by comparing FMA subscores between SCS OFF at baseline and SCS ON at week 4, when effective improvement was maximal. Changes in subscores for each participant were normalized by the maximum score of each subcomponent. For example, an increase of 3 points in ES was normalized by 6, the maximum score for ES, yielding a normalized value of 0.5. These normalized subscores were then summed across all participants for analysis. Note that to assess the effects of SCS on abnormal synergies across joints, we focused on the MCS and MOS subcomponents of the FMA^33–35^. specifically evaluate the ability to perform individualized multi-joint movements outside of the pathological coupling patterns observed after stroke^33–35^, such as extending the elbow while simultaneously flexing the shoulder.

#### Fugl-Meyer Assessment (FMA) for upper-limb sensory function

We used the sensory component of the Fugl-Meyer Assessment (FMA) to quantify proprioception and tactile functions in the upper-extremity. The sensory component of the FMA is assessed using the same score scale as the motor FMA components, with four items for proprioception and two items for tactile functions (maximum score of 12 points). A trained occupational therapist (A.B.) conducted all assessments after administering the FMA for motor function. That is, sensory FMA was administered at the same timepoints and under the same conditions as the motor FMA. Participants were instructed to keep their eyes closed throughout the evaluation. For proprioception testing, the therapist moved the participant’s affected arm/hand, and asked them to mirror the movement with the non-affected arm/hand. For tactile testing, the therapist lightly touched the participant’s affected arm/hand using a piece of cotton, and asked them to verbally identify where touch was felt, if touch was perceived. The FMA sensory scores used in this study were assessed from all participants before the implantation of the SCS lead electrodes (i.e., baseline).

#### Modified Ashworth Scale (MAS)

We used the Modified Ashworth Scale (MAS) to assess both the effects of SCS on upper-limb spasticity. The MAS is a validated clinical tool with good test-retest reliability^38^, used to rate resistance to passive movement on a scale from 0 (no increased muscle tone) to 5 (marked rigidity). Of note, we converted the original +1 score to 2 and rescaled the [2, 4] score range to [3, 5] for comparisons between our results and MCID values^39^. To minimize confounding effects of SCS and physical activity, the MAS was assessed by a trained occupational therapist (A.B.) at the beginning of each experimental session. For each participant, we limited MAS assessments to joints with pronounced spasticity to reduce daily testing time. Overall, the MAS was assessed for shoulder external and internal rotators; shoulder abductors; shoulder flexors; elbow flexors and extensors; wrist flexors, supinators, and pronators; and finger flexors.

Assistive effects of SCS on spasticity were assessed by comparing MAS scores between SCS ON and SCS OFF conditions within the same experimental session. Both conditions were conducted at the beginning of the session. The stimulation conditions were pseudorandomized and blinded to the evaluator. These assistive effects were assessed at week 2 for SCS02 and SCS07; at week 3 for SCS04 and SCS08; at week 4 for SCS01 and SCS05; and they were not assessed for SCS03. Therapeutic effects emerging over time on spasticity were quantified by comparing the average of the MAS scores at week 4 to baseline. A minimal clinically important difference (MCID) of 0.76 was used to interpret meaningful change^39^.

### Surface electromyography (EMG) activity during reach-and-pull task

#### EMG recordings

To assess muscle activity during movement, surface electromyography (EMG) was recorded using a wireless EMG system (Trigno, Delsys Inc.). Up to 12 synchronized wireless sensors (Avanti Trigno, Delsys Inc.) were used to amplify, digitize, and wirelessly transmit EMG signals to a base station unit. Each sensor sampled the analog signal at 1925.925 Hz and applied a hardware bandpass filter of 20-800 Hz. Once the signals were received by the base station, they were converted back to an analog waveform and resampled at 8000 Hz by a data acquisition system (PCI-6255, National Instruments) for synchronization with other task events. The Trigno system has a known, fixed wireless latency of 59.6 ms, which was considered for processing the EMG signals.

At the beginning of each experimental session, the arms and hands were cleaned using mildly abrasive skin preparation gel and isopropyl alcohol. Skin safe adhesive was used to secure the EMG sensors to the subject’s arm. Depending on the muscles of interest for a particular experiment, we recorded from up to 12 individual muscles, such as trapezius, anterior deltoid, biceps brachii, triceps brachii and abductor digiti minimi. Sensors were then carefully removed at the end of each session.

#### Myoelectric activity during planar reach task

To analyze the effect of SCS on muscle activity patterns during the planar reach-and-pull task, we used EMG signals acquired from triceps brachii, biceps brachii and brachioradialis^64,65^. All EMG signals were bandpass-filtered between 25 and 500 Hz using a zero-lag 5th order Butterworth digital filter. We first segmented the EMG signals into reach and pull phases. We then calculated the root-mean square (RMS) of the EMG envelope (eEMG) across SCS ON and OFF conditions for each muscle and movement phase (reach and pull).

#### Agonist–antagonist muscle activation ratio

We quantified the co-contraction of the triceps brachii (TRI), biceps brachii (BIC) and brachioradialis (BR) muscles during the reach-and-pull task using agonist-antagonist activation ratios: TRI/BR and TB/BIC. For each participant, reach target (or pull start) and stimulation condition (SCS ON and SCS OFF), we averaged the myoelectric activity of each muscle—measured as the RMS of the EMG envelope (eEMG)—and the corresponding agonist–antagonist activation ratios. Finally, we quantified changes in muscle co-contraction due to SCS as the percentage change in agonist-antagonist activation ratios, computed as the ratio of values during SCS ON relative to SCS OFF (i.e., SCS ON/SCS OFF):

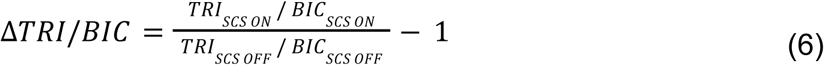

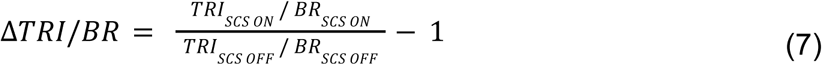

### Time on active movement

Although a formal rehabilitation protocol was not implemented, participants engaged in active movements during task assessments across study. We estimated the time spent on active movement under SCS ON and OFF conditions using video recordings from the experimental sessions and the length of data files. Task assessments included strength tasks using the HUMAC isokinetic system, reaching tasks using the KINARM exoskeleton robot, and other functional tasks. Break intervals were not included in the time calculations.

### Statistics

#### Bootstrapping

For independent comparisons at subject-level, we performed a bootstrap analysis in which each condition was resampled separately, and the distribution of the difference in means was used to evaluate condition-level effects. For example, SCS ON versus SCS OFF conditions were compared for shoulder torque in participant SCS03 (**Fig. 2b**). For paired comparisons at the group level, we performed a bootstrap analysis by resampling the paired differences across participants and testing whether the distribution of these differences was significantly different from zero. For example, FMA scores compared between SCS ON and SCS OFF conditions across *n* = 7 participants in **Fig. 4c**. In both analyses, 10,000 bootstrap samples were used to construct empirical confidence intervals (CIs) for quantities of interest. A 95% CI was obtained by identifying the 2.5th and 97.5th quantiles for the resulting values. The null hypothesis of no difference in the mean was rejected if 0 was not included in the 95% CI. Bootstrap statistical analysis was only performed when at least five data points were obtained.

For the total MVC change across joints, we performed the bootstrap analysis by resampling at the subject-level, computing each within-join mean, and then computing the average across joints. Resampling all values for each subject preserves the correlation across joints within a subject.

#### Linear correlations

Pearson correlation analyses were conducted to examine the linear relationship between the percentage change in agonist-antagonist muscle activation ratios (TRI/BIC and TRI/BR) and the corresponding percentage changes in arm kinematic metrics (path efficiency, deviation error, velocity peaks and log dimensionless jerk) enabled by SCS. Average values for the change in agonist-antagonist muscle activation ratios and arm kinematics were pooled across all participants and reach/pull targets. To reduce bias of extreme values, data points outside 1.5 times the interquartile range (IQR) of the distribution of myoelectric activity change across stimulation conditions (SCS ON/SCS OFF) for TRI, BIC, or BR were excluded from the analysis. All correlation coefficients and p-values were derived using SPSS Statistics (IBM, SPSS Inc.). Statistical significance set at α = 0.05.

## Data Availability

All data produced in the present study are available upon reasonable request to the authors.

## ACKNOWLEDGEMENTS

We thank T. Simpson for engineering support. The study was executed through the support of National Institutes of Health Brain Initiative grant no. UG3NS123135-01A1 to M.C. and D.J.W. and internal funding from the Department of Neurological Surgery at the University of Pittsburgh to M.C., the Department of Mechanical Engineering and the Neuroscience Institute at Carnegie Mellon University to D.J.W. and the Department of Physical Medicine and Rehabilitation at the University of Pittsburgh to E.P.

## ETHICS DECLARATIONS

M.P.P, D.J.W., M.C. and P.C.G. are founders and shareholders of Reach Neuro, a company developing spinal cord stimulation technologies for stroke. E.P. has interest in Reach Neuro due to personal relationships with M.C.

M.P.P, D.J.W., M.C., P.C.G., E.P., E.S., N.V. and E.C. are inventors on several patents related to this work. All other authors declare no competing interests.

## CONTRIBUTIONS

M.C., D.J.W. and E.P. conceived the study. M.C., D.J.W. and E.P. secured the funding. M.C., E.P., J.W.K. and D.J.W. designed the experiments and assessments. M.P.P., D.J.W., N.V. and E.S. designed and implemented the stimulation control system. E.C. and E.P. designed and implemented the KINARM motor tasks. S.E., R.M.dF. and E.P. designed and performed the MRI-based analyses. A.B., R.M.dF and G.F.W. implemented patient recruitment, eligibility and monitoring, and coordinated management of the study. M.C., L.E.F. and P.C.G. designed the neurosurgical approach. P.C.G., D.P.F. and R.M.F. implemented and evaluated the neurosurgical procedures and participants’ clinical management protocols. J.B., R.M.dF, M.P.P., S.G and M.C. designed and performed the intraoperative monitoring protocols. R.M.dF., M.P.P., N.V., E.S., E.C., L.B., M.C., E.P., A.B. and J.W.K. performed the experiments. J.G. and R.M.dF designed the statistical data analyses. R.M.dF., S.B., L.B., M.C., J.W.K. and E.P. interpreted the results. R.M.dF. and S.B. analyzed the data. R.M.dF. and S.B. created the figures. M.C., R.M.dF. and S.B. wrote the paper and all authors contributed to its editing.

## EXTENDED DATA

**Table 1:**
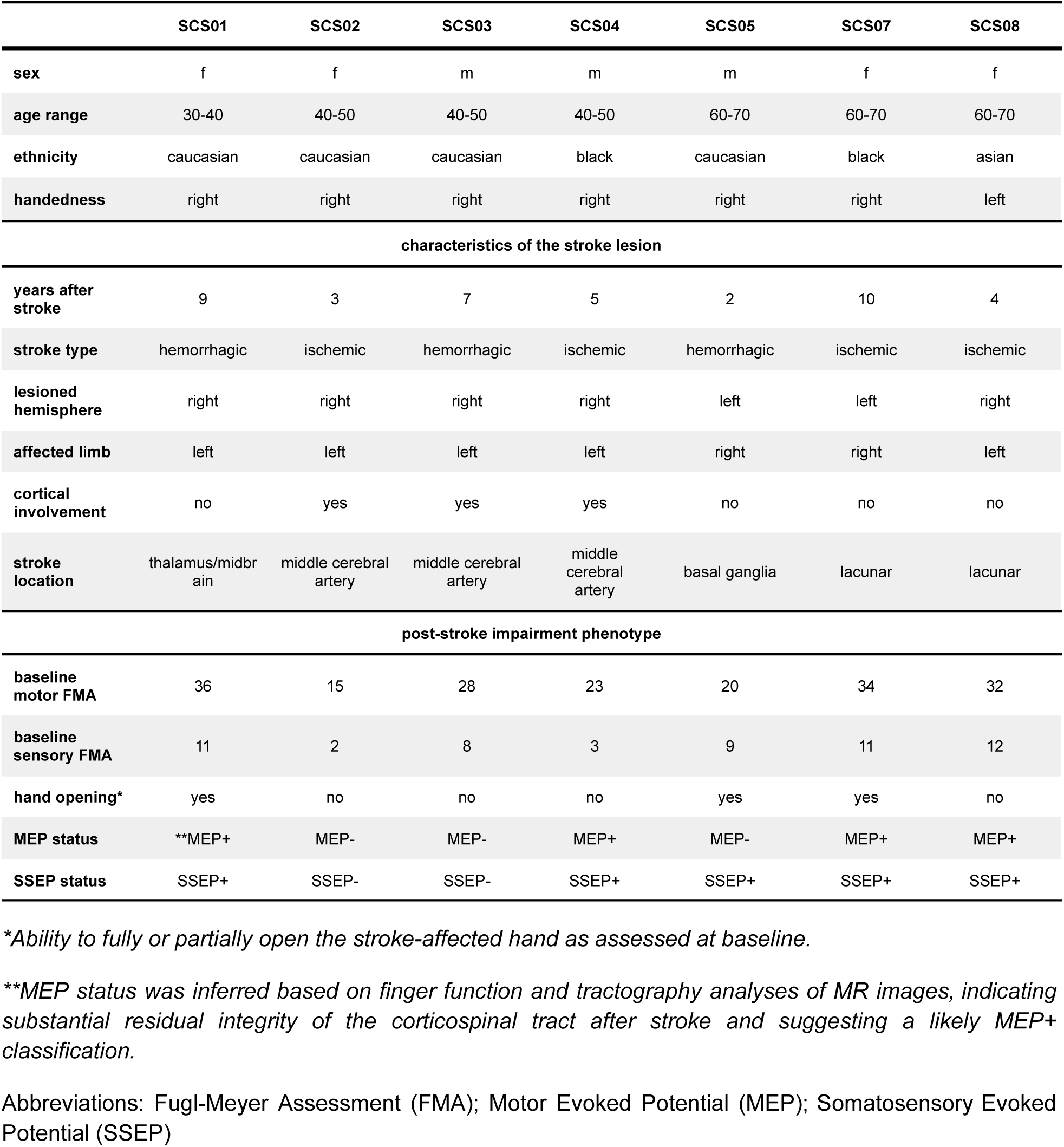
Demographics of the stroke participants in the clinical trial. Characteristics of the stroke lesion and pos-stroke impairment phenotype description of all participants in the clinical trial.

**Table 2:** Details of adverse events. Detailed description and resolution of all adverse events occurred during the clinical trial. No serious adverse events occurred.

| participant | severity | relationship with the study | description | resolution |
| --- | --- | --- | --- | --- |
| SCS01 | 1. mild | 1. related | 1. pain at the site where the leads exit the body; | 1. pain was resolved by replacing the sutures at the leads exit site; |
|  | 2. mild | 2. possibly related | 2. phlebitis of left (affected) arm. | 2. phlebitis was resolved with antibiotics; |
| SCS02 | 1. mild | 1. possibly related | 1. bruise on the back of the left (affected) arm. | 1. resolved spontaneously within a week. |
| SCS03 | - | - | - | - |
| SCS04 | 1. mild | 1. possibly related | 1. persistent hiccups; | 1. resolved spontaneously within a week; |
|  | 2. mild | 2. possibly related | 2. pain in the left (affected) arm and leg; | 2. resolved spontaneously within a week; |
|  | 3. mild | 3. related | 3. difficulty in breathing when stimulated at high amplitude (i.e., 5 mA) with rostral contacts located between C3 and C4 spinal levels. | 3. stimulation was immediately discontinued, resolving the respiratory issue. |
| SCS05 | 1. mild | 1. possibly related | 1. headache followed by experimental session; | 1. resolved spontaneously within one day; |
|  | 2. mild | 2. related | 2. mediolateral lead migration. | 2. electrode migration did not cause negative effects on motor functions. |
| SCS07 | 1. mild | 1. possibly related | 1. vomit prior experimental session; | 1. resolved spontaneously within one day; |
|  | 2. mild | 2. possibly related | 2. vomit during experimental session; | 2. participant was assessed by the study physician (G.W.) and the experimental session was discontinued. The adverse event was resolved spontaneously within one day; |
|  | 3. mild | 3. not related | 3. participant fell from her scooter while commuting to experimental space; | 3. participant had no concerning injuries; |
|  | 4. mild | 4. possibly related | 4. pain and pinching sensation to upper trapezius area of (affected) right arm; | 4. resolved spontaneously within a couple of days; |
|  | 5. mild | 5. possibly related | 5. swelling around the scapular region with sensitivity to touch; | 5. resolved spontaneously within a couple of days; |
|  | 6. mild | 6. not related | 6. drowsiness due to high blood pressure before the experimental session. | 6. participant was assessed by the study physician (G.W.) and the experimental session was discontinued. The adverse event was resolved spontaneously within one day; |
| SCS08 | - | - | - | - |

**Table 3:** Modified Ashworth Scale (MAS) scores for each participant. Assistive of spinal cord stimulation (SCS) and therapeutic effects emerged over time on MAS scores across participants and muscle groups.

| muscle group | SCS01 | SCS02 | SCS03 | SCS04 | SCS05 | SCS07 | SCS08 | AVERAGE |
| --- | --- | --- | --- | --- | --- | --- | --- | --- |
| **assistive effects (SCS ON - SCS OFF)** |  |  |  |  |  |  |  |  |
| shoulder external rotators | 0.00 | 0.00 |  | 0.00 |  | 0.00 | 0.00 | **0.00** |
| shoulder internal rotators | 0.00 | 1.00 |  | 0.00 | 0.00 | 0.00 | 0.00 | **0.17** |
| shoulder abductors | 0.00 | 1.00 |  |  |  |  |  | **0.50** |
| shoulder flexors |  |  |  | 0.00 |  |  |  | **0.00** |
| elbow flexors | 0.00 | 0.00 |  | -1.00 | 0.00 | -1.00 | 0.00 | **-0.30** |
| elbow extensors |  | 0.00 |  | 0.00 | -1.00 | 0.00 | 0.00 | **-0.20** |
| wrist flexors |  | 0.00 |  | 0.00 | -1.00 | -1.00 | -1.00 | **-0.60** |
| wrist supinators | 0.00 |  |  | 0.00 |  | -1.00 | 0.00 | **-0.30** |
| wrist pronators |  | 0.00 |  | 0.00 | 0.00 | -1.00 | 0.00 | **-0.20** |
| fingers flexors | 0.00 | -1.00 |  | 0.00 | 0.00 | -1.00 | 0.00 | **-0.30** |
| **TOTAL** | **0** | **1.00** |  | **-1.00** | **-2.00** | **-5.00** | **-1.00** | **-1.30** |
| **therapeutic effects (WEEK 4 - BASELINE under SCS OFF)** |  |  |  |  |  |  |  |  |
| shoulder external rotators | -0.33 | -1.0 |  |  |  | -1.50 | 0.00 | **-0.71** |
| shoulder internal rotators | -0.33 | -0.50 | -1.50 |  | -1.67 | 1.00 | 0.00 | **-0.50** |
| shoulder abductors | -0.33 | -2.25 | -3.00 |  |  |  |  | **-1.86** |
| elbow flexors | -1.00 | -0.25 | -0.75 | 0.00 | 0.00 | 0.00 | -1.00 | **-0.43** |
| elbow extensors |  |  | 0.00 |  | 0.33 | -1.50 | 0.00 | **-0.29** |
| wrist flexors |  | 0.00 | -0.25 | -1.00 | 1.33 | -0.25 | 0.00 | **-0.03** |
| wrist supinators | -0.33 |  |  |  |  | -1.50 | 1.00 | **-0.28** |
| wrist pronators |  |  | -2.25 |  | -1.00 | -0.50 | 0.50 | **-0.81** |
| fingers flexors | -0.33 | -1.5 | -1.75 | -1.00 | -0.33 | 0.25 | 0.00 | **-0.66** |
| **TOTAL** | **-2.65** | **-5.50** | **-9.50** | **-2.00** | **-1.34** | **-4.00** | **0.50** | **-5.57** |

**Extended Data Figure 1:**
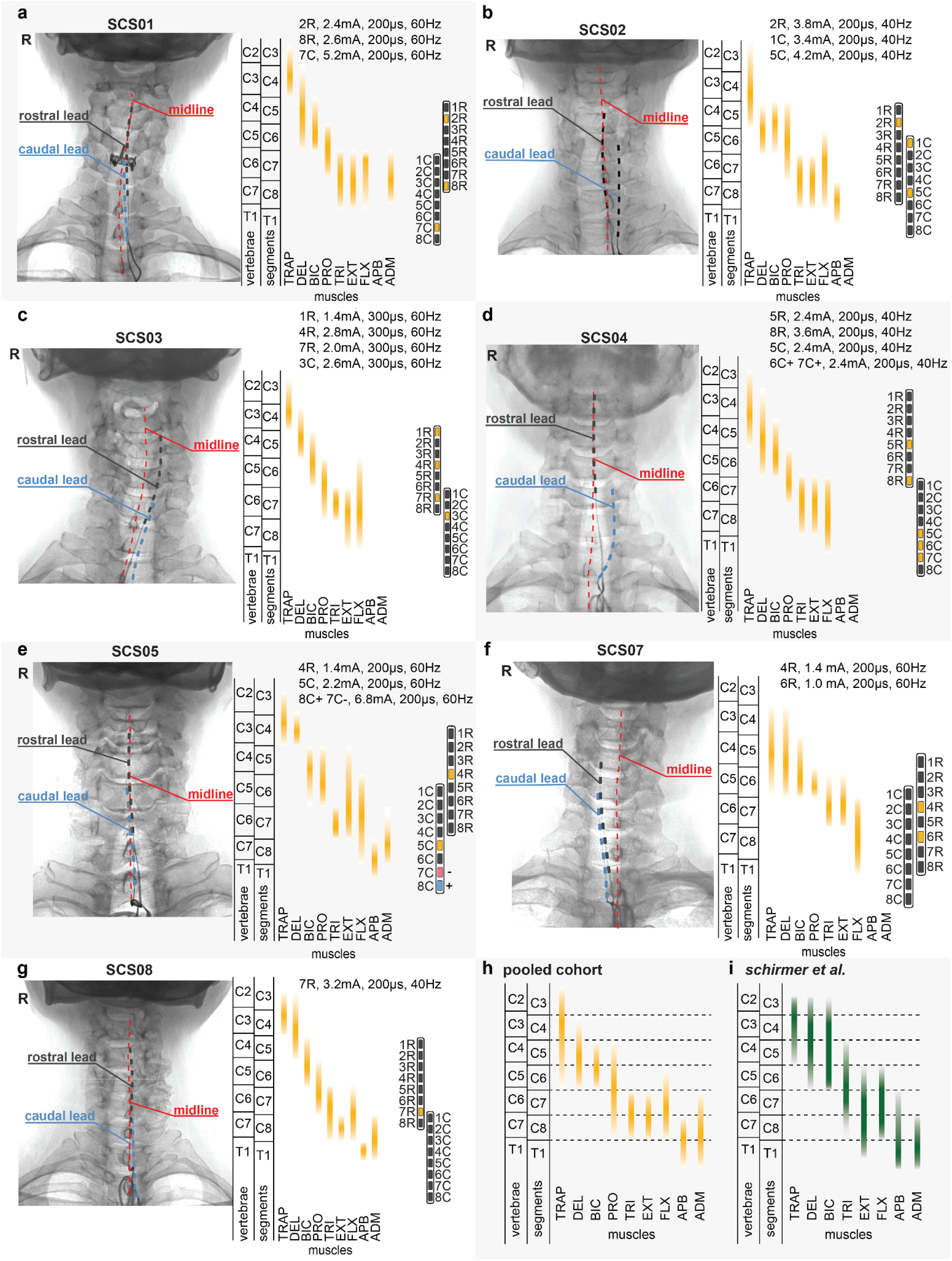
Spinal segment maps. **a-g.** SCS lead placement, spinal segment maps, and clinical configuration for each participant. X-rays taken at week 2 show the rostral (black) and caudal (blue) SCS leads, lateralized relative to the spinal midline (dashed red trace). Spinal segment maps, estimated using intraoperative neurophysiological recordings, show the distribution of motor neuron pools across cervical spinal levels. Spinal segment maps were used to calibrate the SCS parameters for each participant. Contacts filled in yellow indicate monopolar configurations, while contacts filled in blue and red indicate bipolar configurations. **h**. Pooled spinal segment map of the motoneuron pools from the cohort of seven participants. **i.** Reference spinal segment map of motor neuron pools adapted from Schirmer et al.^30^.

**Extended Data Figure 2:**
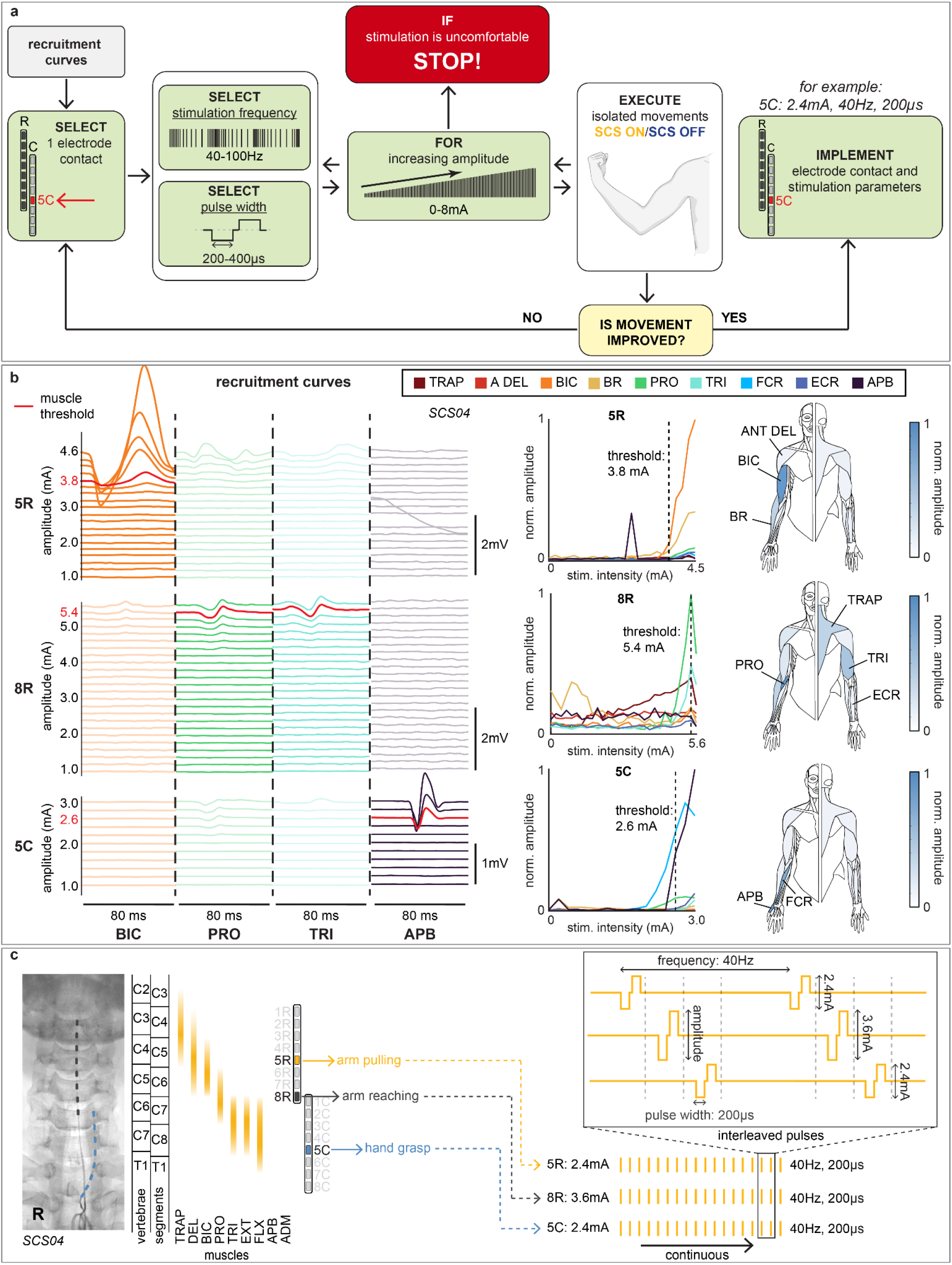
Calibration of spinal cord stimulation (SCS) parameters. **a.** Systematic procedure for calibration of SCS parameters. Recruitment curves are used to identify electrode contacts for calibration. Pulse width, stimulation frequency and amplitude are then interactively adjusted while assessing the assistive effects of SCS on movement performance (i.e., difference between SCS ON and OFF conditions). **b.** Representative recruitment curves of electrode contacts 5R, 8R and 8C for SCS04. Left: Contacts 5R, 8R and 8C selectively targeted activation of biceps brachii (BIC), pronator teres (PRO), triceps brachii (TRI), and abductor pollicis brevis (APB) muscles. For each contact, motor threshold was defined as the minimum stimulation amplitude necessary to evoke an electromyography (EMG) response with peak-to-peak ≥50 µV (red trace). Middle: recruitment curves, normalized to the maximum peak-to-peak response of each muscle, show the activation of proximal and distal muscles for each contact. Abbreviations: Trapezius (TRAP); Anterior Deltoid (A DEL); brachioradialis (BR); flexor carpi radialis (FCR); extensor carpi radialis (ECR). Right: human drawings summarize the activation profile associated with each recruitment curve. **c.** Example of calibrated SCS parameters for SCS04. Based on the recruitment curves in panel b, contacts 5R, 8R and 8C were calibrated to facilitate arm pulling, arm reaching and hand grasping.

**Extended Data Figure 3:**
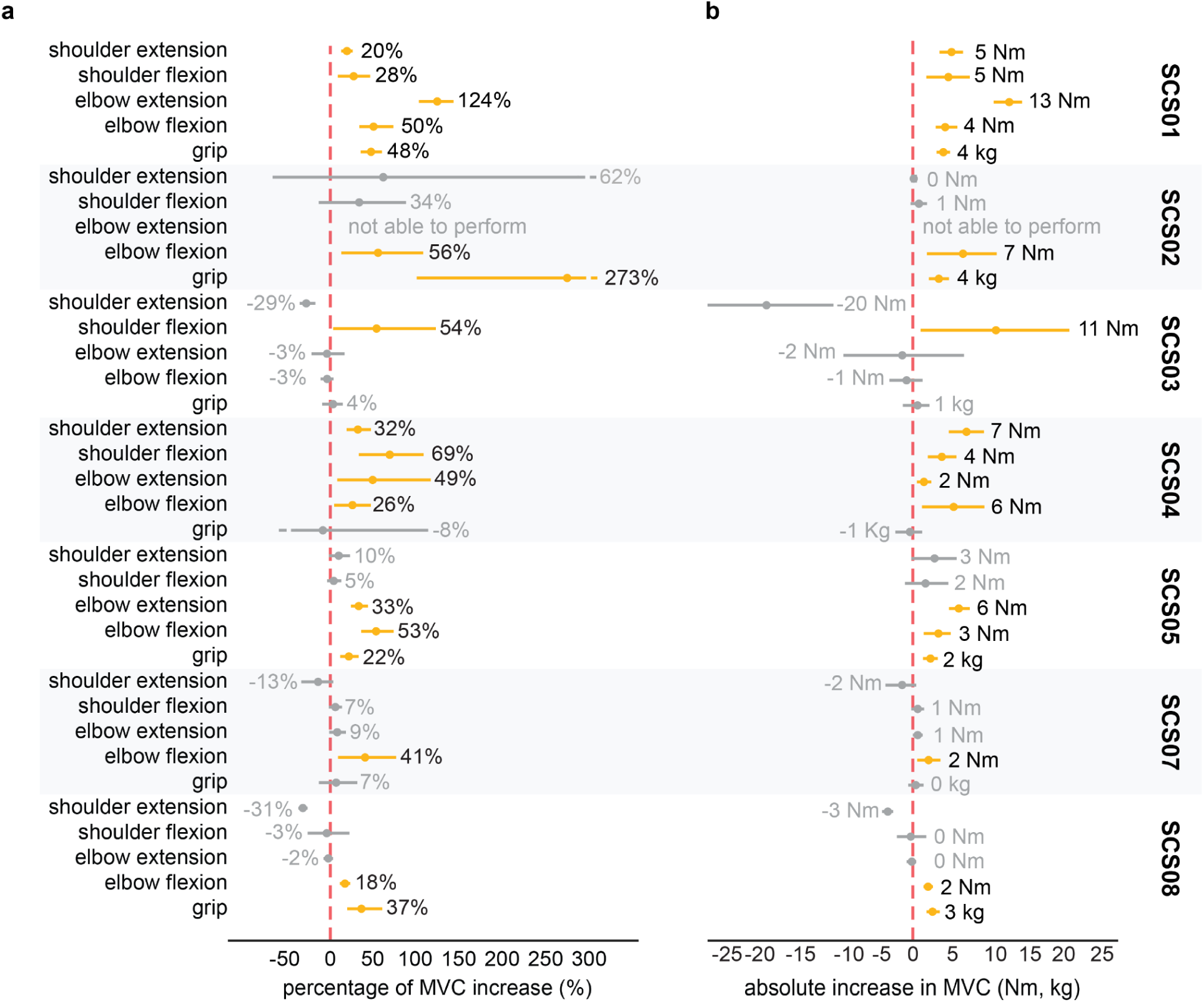
MVC values for each participant. **a.** Maximum voluntary contraction (MVC) strength was measured for five isometric joint movements: shoulder extension, shoulder flexion, elbow extension, elbow flexion, and grip. Assistive effects of spinal cord stimulation (SCS) on strength were quantified by the average difference in MVC force between SCS ON and OFF conditions. Each participant showed significant improvement in at least one joint. Horizontal lines in yellow highlight significant improvements in strength enabled by SCS. **b.** Absolute increase in MVC values between SCS OFF to SCS ON across all joints and all participants.

**Extended Data Figure 4:**
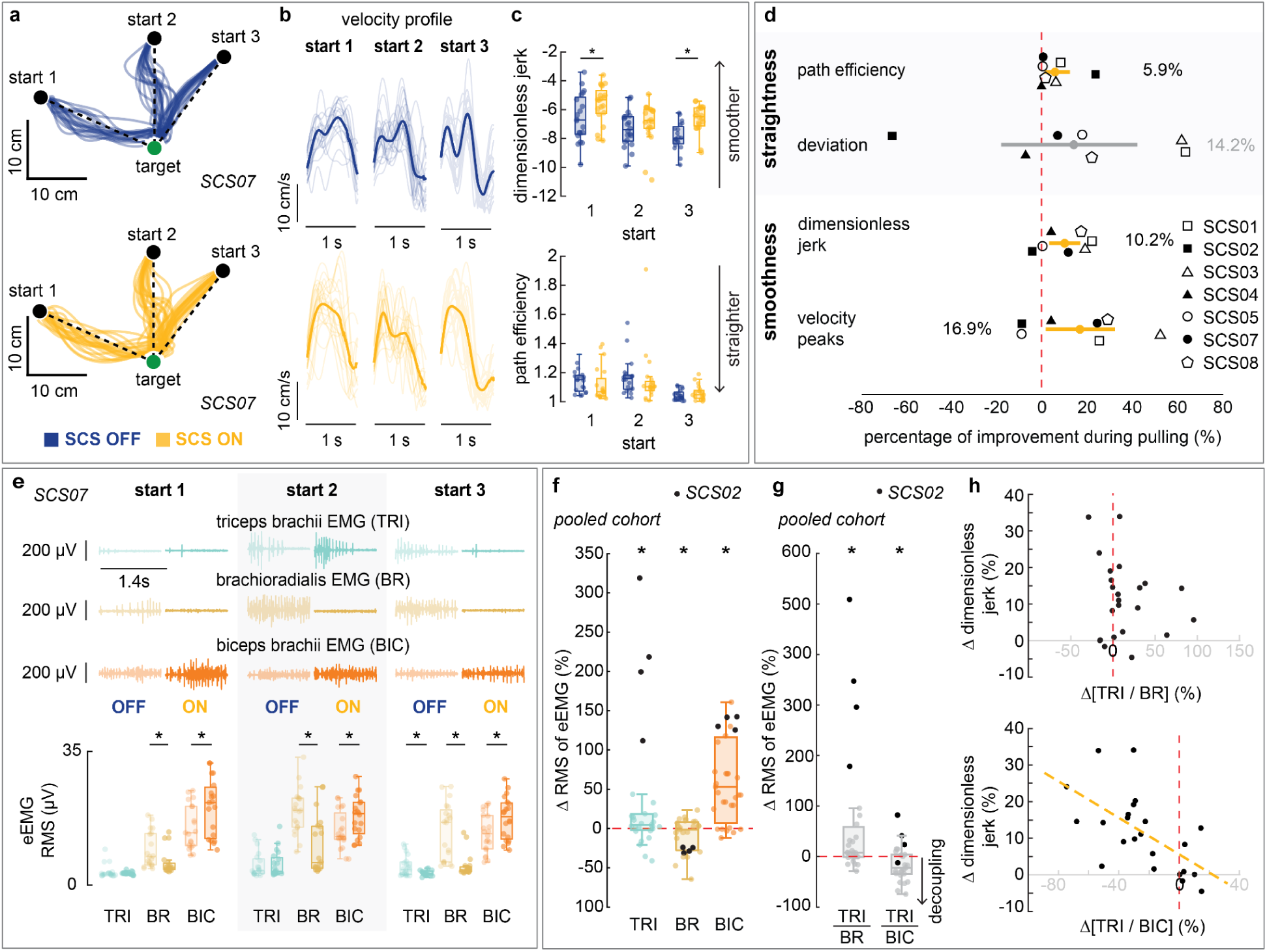
Assistive effects of SCS on arm pull kinematics (dexterity). **a-c**. Examples from participant SCS07 showing smoother pull trajectories during SCS ON. A planar reach-and-pull task was used to assess the assistive effects of spinal cord stimulation (SCS) on upper-limb dexterity. During the pull phase, participants were instructed to return the hand to the initial position from a reached target. **a.** The geometry of pull trajectories are not qualitatively different between SCS ON and OFF. **b.** Hand velocity profiles corresponding to the trajectories in panel a. show less variability during SCS ON compared to SCS OFF. Traces in darker shades indicate the average across repetitions. **c.** Consistent with panel b., trajectory smoothness, measured as log dimensionless jerk, is significantly improved during SCS ON, while trajectory straightness, measured by path efficiency, did not change during SCS ON. **d.** Assistive effects of SCS on arm pull kinematics pooled across all participants. We observed significant improvements in all metrics except for the deviation error. **e.** Electromyography (EMG) recordings during pull trajectories in panel a., for SCS07, show increased activation of biceps brachii (BIC), and reduced activity in brachioradialis (BR) and triceps brachii (TRI) during SCS ON (darker shades) compared to SCS OFF (lighter shades). Root-mean-square (RMS) of the EMG envelope (eEMG) was used to quantify muscle activity across each target. **f-h**. Assistive effects of SCS on muscle synergies during the pull phase. Asterisks in panels f and g indicate statistically significant differences between SCS ON and OFF conditions. **f.** Percentage change in RMS of eEMG signals from TRI, BR and BIC muscles between SCS ON and OFF conditions, pooled across all participants. Notably, TRI muscle activity increased significantly during SCS ON, largely driven by outlier values from SCS02 (black dots). **g.** Agonist-antagonist activation ratios (TRI/BR and TRI/BIC) were used to quantify changes in muscle co-contractions between SCS ON and OFF conditions (SCS ON/ SCS OFF -1). A negative ratio (< 0) indicates decoupling of the agonist-antagonist muscles. The agonist-antagonist ratio TRI/BIC was significantly increased with SCS ON, whereas TRI/BR was not statistically different across stimulation conditions. Outlier values from SCS02 are marked with black dots. **h.** Correlation between the percentage change in agonist-antagonist muscle activation ratios (TRI/BIC and TRI/BR) and the corresponding percentage changes in log dimensionless jerk enabled by SCS during the pull phase. A significant negative linear correlation was observed for TRI/BIC (r=-0.612; p<0.001), indicating reduced co-contraction between the TRI and BIC under SCS ON was associated with improved movement smoothness. No significant correlation was found for TRI/BR (r=-0.195; p=0.186).

**Extended Data Figure 5:**
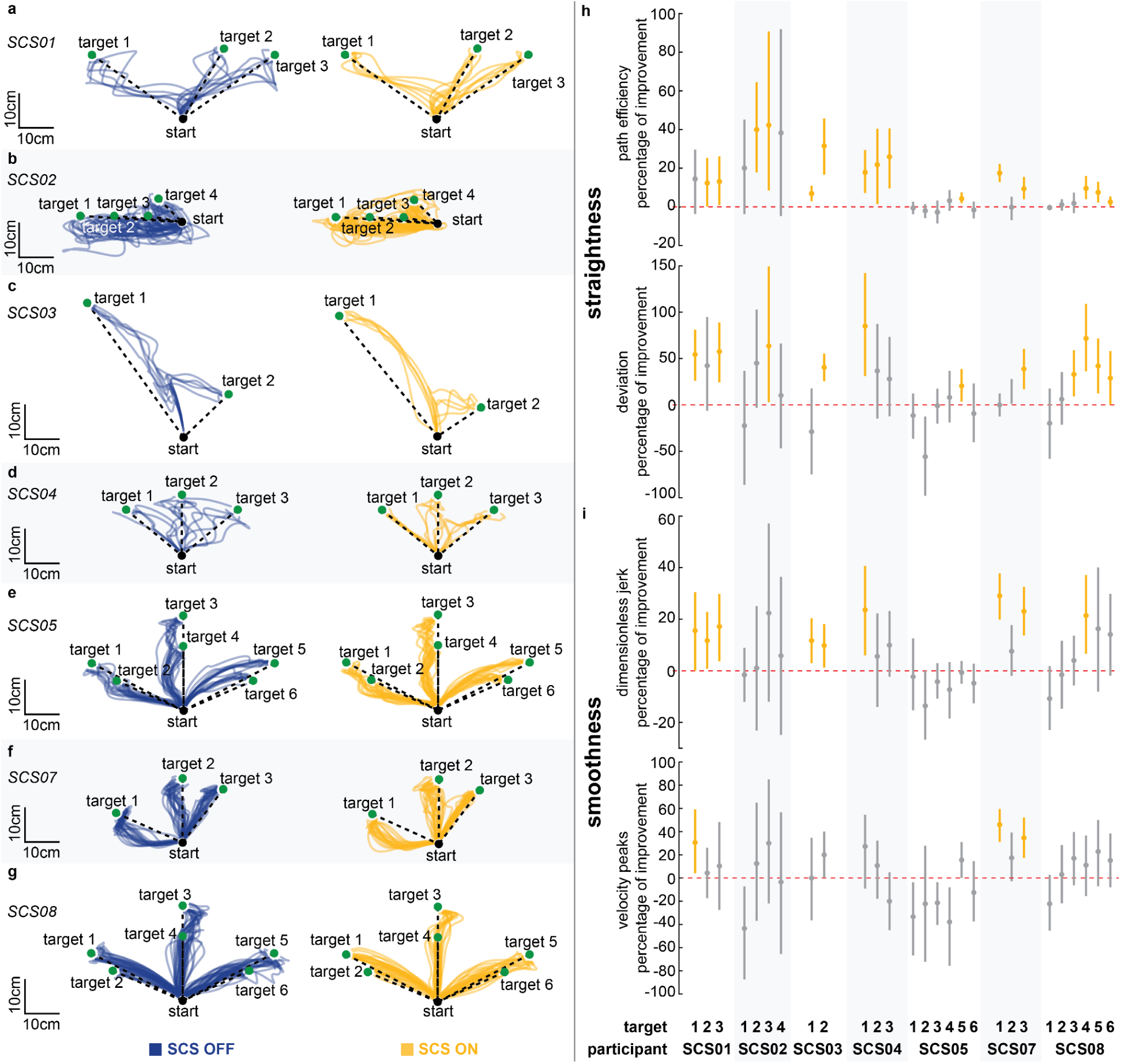
Arm kinematics during the reach phase for each participant. **a-g.** Reach trajectories of each participant (SCS01-SCS08) under spinal cord stimulation (SCS) OFF (blue) and SCS ON (yellow) conditions. **h,i.** Arm kinematic metrics quantifying straightness **(h)** and smoothness **(i)** of the trajectories shown in panels a-g for each target position. Confidence intervals (5%–95%) of the percentage difference in kinematic metrics between SCS ON and OFF conditions are displayed with horizontal lines around the average (circle). Horizontal traces highlighted in yellow indicate targets with significant improvements in arm kinematics during the pull phase.

**Extended Data Figure 6:**
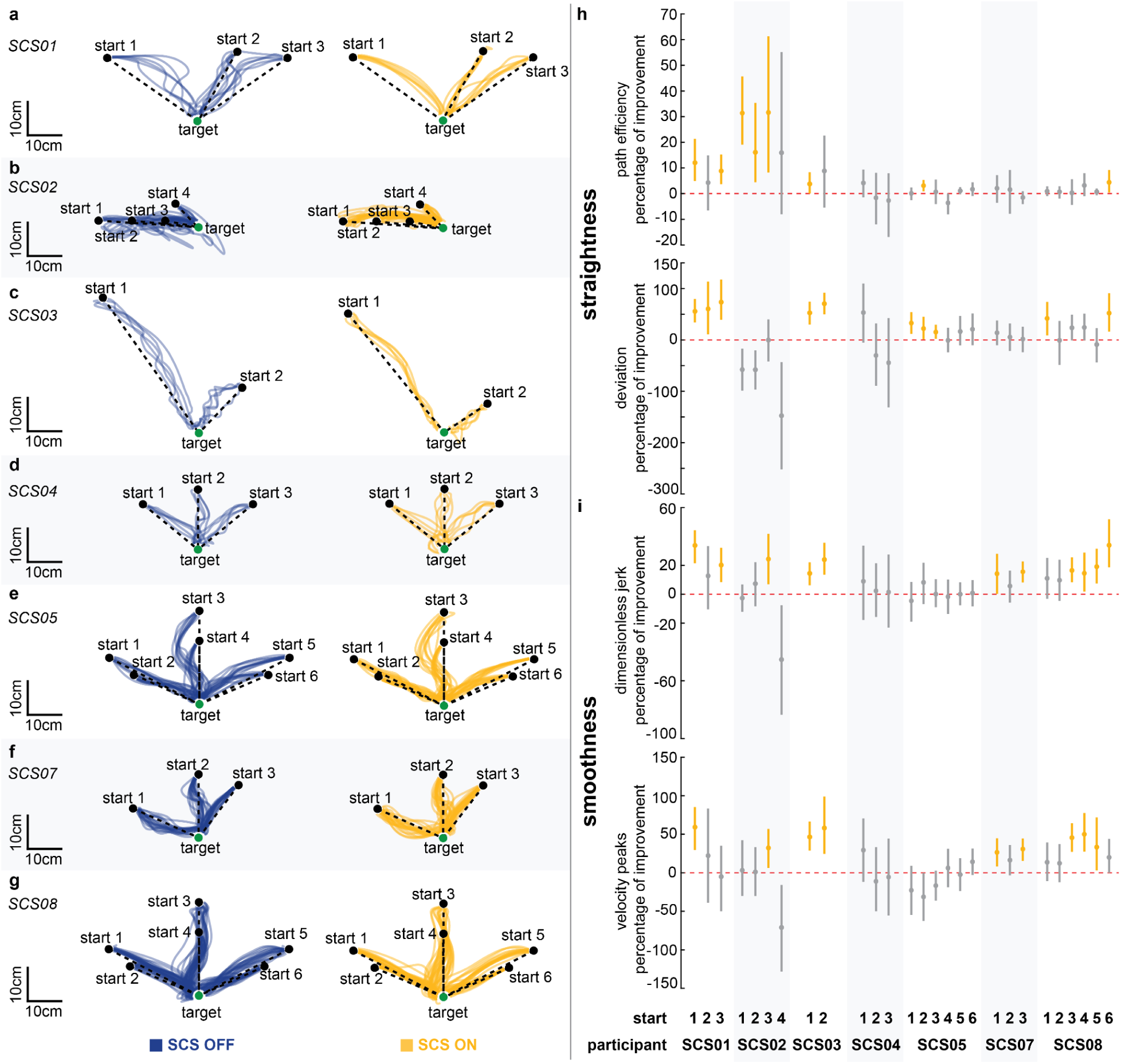
Arm kinematics during the pull phase for each participant. **a-g.** Pull trajectories of each participant (SCS01-SCS08) under spinal cord stimulation (SCS) OFF (blue) and SCS ON (yellow) conditions. **h,i.** Arm kinematic metrics quantifying straightness **(h)** and smoothness **(i)** of the trajectories shown in panels a-g for each start position. Confidence intervals (5%–95%) of the percentage difference in kinematic metrics between SCS ON and OFF conditions are displayed with horizontal lines around the average (circle). Horizontal traces highlighted in yellow indicate targets with significant improvements in arm kinematics during the pull phase.

**Extended Data Figure 7:**
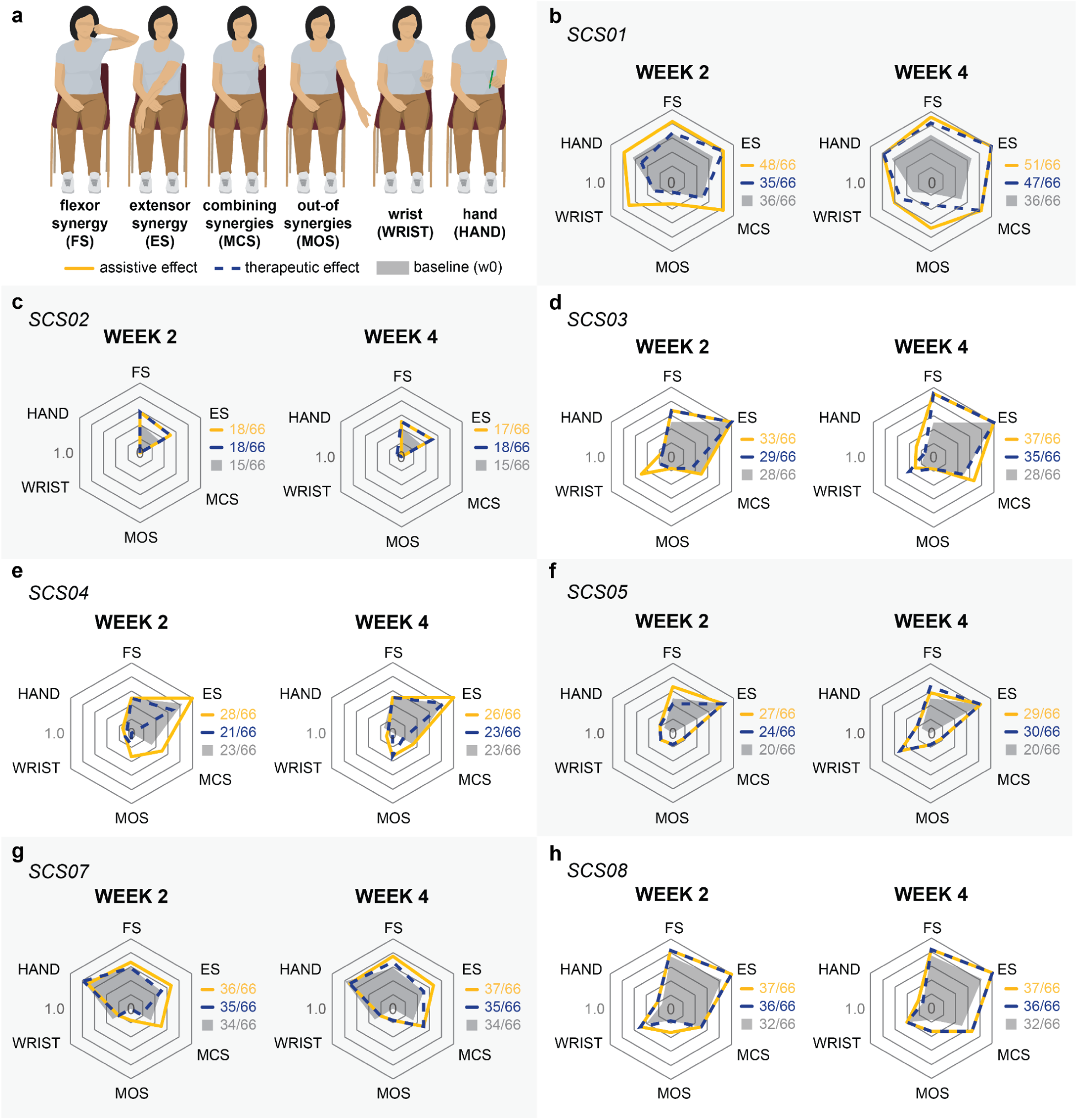
Fugl-Meyer Assessment (FMA) subscores for each participant. **a.** The FMA for upper-extremity motor function was used to assess the effects of SCS. **b-h.** For each participant, web plots show the normalized change in scores of FMA subcomponents during SCS ON and OFF conditions across different timepoints. The FMA was assessed at baseline (pre-implant), week 2 and week 4. Assistive effects refer to the change in FMA scores between SCS ON and OFF conditions at a given timepoint (week 2 or week 4), whereas therapeutic effects refer to the change in FMA scores over time for the SCS OFF condition. Baseline FMA scores are shaded in grey, assistive effects in yellow, and therapeutic effects in dashed blue.

