## Supplementary Information for "SPINAL CORD STIMULATION IMPROVES MOTOR FUNCTION AND SPASTICITY IN CHRONIC POST-STROKE UPPER LIMB HEMIPARESIS"

#### METHODS

##### Surgical Procedure

To implant a pair of spinal cord stimulation (SCS) leads, general anesthesia was induced using propofol and maintained using total intravenous anesthesia (TIVA) at levels that allowed for reliable somatosensory evoked potential monitoring. To facilitate intraoperative monitoring of SCS-evoked electromyographic (EMG) responses, short-acting paralytic was administered only during intubation. Participants were positioned prone and secured using a 3-pin Mayfield head holder. The back and neck were then prepped and draped in a standard sterile fashion. Prophylactic antibiotics were administered. A small incision over the T1-T2 laminas was performed to expose the fascia. A pair of clinically approved 8-contact percutaneous spinal leads (PN 977A260, Medtronic) were placed through a Tuohy needle inserted into the T1-T2 epidural interspace. The rostral lead was inserted first. The rostral lead was steered in situ along the lateral aspect of the spinal cord, such that the most distal contact was positioned at the base of the C3 vertebral process. The caudal lead was placed to span the lower cervical spinal levels down to the T1 vertebral process, overlapping partially with the rostral lead (**Extended Data Fig. 1**). Fluoroscopy was used to guide lead positioning. Appropriate lead placement was verified by testing frequency dependent suppression of SCS evoked EMG responses. An intraoperative neuromonitoring system (Xltek Protektor, Natus Medical) was used to deliver stimulation and record evoked EMG responses bilaterally in both arms. Suppression of EMG evoked responses were observed by delivering SCS at 20Hz with amplitude above motor threshold. The Tuohy needle was removed after each lead was placed. The proximal ends of the leads were sutured to the fascia to prevent migration. The distal ends were tunneled subcutaneously and brought out through a separate stab incision over the ipsilateral flank. Both incisions were closed, leaving the externalized portion of the leads covered.

The leads were explanted four weeks after implantation. Participants underwent a preparation procedure similar to the implantation surgery. The cervical incision at the upper thoracic region was reopened to access the leads, which were cut and removed. The distal ends were removed through the lateral incision. Both cervical and lateral incisions were then closed.

#### Custom Stimulation Controller

During the trial, a custom stimulation controller was used to deliver SCS during experimental sessions. This custom stimulation system interfaced the external leads with a single channel, current controlled stimulator (DS8R, Digitimer) connected to a 1-to-8 channel multiplexer. A programmed microcontroller board (Arduino Due, Arduino), controlled through a custom-built GUI, was used to configure the stimulation amplitude and frequency delivered at each contact electrode. Stimulation pulses had cathodic-first, biphasic, symmetric and square waveforms with 200 us to 400 us pulse width and 10 us to 50 us interphase interval. More details about the custom stimulation controller can be found in Powell et al., 2023.

#### Action Research Arm Test (ARAT)

The Action Research Arm Test (ARAT) is an assessment of upper-limb motor function that focuses on object interaction and manipulation. It consists of 19 task items scored from 0 to 3 (maximum score is 57), where 0 indicates the participant is unable to perform the task and 3 indicates the participant can perform it normally. These 19 task items are distributed across four categories: grasp, pinch, grip and gross movement.

The ARAT was administered at baseline (pre-implant), week 4 post-implant, and follow-up (a month post-explant) time points by a trained occupational therapist (A.B.). At week 4, the ARAT was conducted under both SCS ON and OFF conditions in pseudorandom order within the same experimental session. The therapist was blinded to the stimulation conditions, and a rest break (5-10 minutes) was given between conditions.

#### Figure Generation

All figures were composed in Adobe Illustrator CC v29.0-29.3.1.

### TABLES

**Supplementary Table 1.** Description of all primary and secondary outcome measures of the clinical trial (clinicaltrials.gov ID NCT04512690). The last column indicates which outcome measures were assessed in this study.

| Outcome Measure | Measure Description | Assessed in Study |
|---|---|---|
| | **primary outcome: safety of epidural cervical spinal cord stimulation (SCS)** | |
| Adverse Events | Study is considered successful if no serious adverse events related to the use of electrical stimulation are reported | Yes |
| Discomfort and Pain | We will assess the relative level of discomfort and/or pain that is associated with the delivery of stimulation to the spinal cord. After each stimulation train, patients will be asked to report their perceived discomfort level using a 10 value subjective scale. Low values will be assigned to low discomfort, and high values to high discomfort.The study is considered successful if 70% of recruited subjects does not report discomfort or pain at stimulation amplitudes that are required to obtain motor responses in the muscles of the arm and hand. | Yes |
| | **secondary outcomes: upper limb motor function** | |
| Motor Impairment | The Fugl-Meyer Assessment (FMA) is a stroke-specific, performance-based impairment index. It is designed to assess motor functioning, balance, sensation and joint functioning in patients with post-stroke hemiplegia. It is applied clinically and in research to determine disease severity, describe motor recovery, and to plan and assess treatment. The upper extremity motor function score ranges from 0 to 66 points. Minimal Detectable Change (MDC) is 5.2 points. The MCID (Minimally Clinically Important Difference) is 4.25 to 7.25. | Yes |
| Dexterity / Function: Action Research Arm Test | The investigators will use the Action Research Arm Test (ARAT) assessment to quantify functional hand and arm dexterity. Performances will be compared with SCS-on against SCS-off. The investigators will consider as a minimally acceptable improvement an increase in the affected arm total score of >4 points. Comparison will be done per patient between Stim-on, Stim-off and pre-study baselines. Maximum score on the test is 57 points, minimum score is zero points, with a higher value indicating better dexterity/function. | Yes |
| Single Joint Force | Isometric torque: measure the isometric torque produced by the subject at the shoulder, elbow and wrist joints. Comparison of SCS-on with SCS-off performance. Success Criteria: ≥20% increased torque production over SCS-off baseline as measured during single-joint isometric torque. | Yes |
| Joint Velocity | The investigators will use the KINARM robot to quantify joint velocity. The investigators will measure 2D kinematics of the arm during several different horizontal reaching tasks. The investigators will also quantify joint velocity in 3D while subjects perform reach and grasp tasks unsupported. Subjects will be tasked to reach targets or objects and manipulate objects while 3D videos of their arm and hand movements are recorded. Arm and hand kinematics will then be analyzed offline in parallel to EMG analysis of arm and hand muscles. Comparison will be done per patient between Stim-on and Stim-off at different time-points. Given the scientific nature of this task no minimal acceptable improvement is defined and data will be used to understand effects of SCS on arm kinematics. | Yes |
| Movement Smoothness | The investigators will use the KINARM robot to quantify movement smoothness. The investigators will measure 2D kinematics of the arm during several different horizontal reaching tasks. The investigators will also quantify movement smoothness in 3D while subjects perform reach and grasp tasks unsupported. Subjects will be tasked to reach targets or objects and manipulate objects while 3D videos of their arm and hand movements are recorded. Arm and hand kinematics will then be analyzed offline in parallel to EMG analysis of arm and hand muscles. Comparison will be done per patient between Stim-on and Stim-off at different time-points. Given the scientific nature of this task no minimal acceptable improvement is defined and data will be used to understand effects of SCS on arm kinematics. | Yes |
| Time to Target | The investigators will use the KINARM robot to quantify time to target. The investigators will measure 2D kinematics of the arm during several different horizontal reaching tasks. The investigators will also quantify time to target in 3D while subjects perform reach and grasp tasks unsupported. Subjects will be tasked to reach targets or objects and manipulate objects while 3D videos of their arm and hand movements are recorded. Arm and hand kinematics will then be analyzed offline in parallel to EMG analysis of arm and hand muscles. Comparison will be done per patient between Stim-on and Stim-off | Yes |

| Outcome Measure | Measure Description | Assessed in Study |
|---|---|---|
| | at different time-points. Given the scientific nature of this task no minimal acceptable improvement is defined and data will be used to understand effects of SCS on arm kinematics. | |
| Spasticity | The investigators will quantify spasticity scores using the Modified Ashworth Scale (MAS) for the shoulder, elbow and wrist joint and compare values with SCS-on and SCS-off. The investigators will consider as a minimally acceptable improvement a decrease of MAS >1, if available for the specific joint. Comparison will be done per patient between Stim-on and Stim-off and pre-study baselines. Maximum score on the MAS is 4, minimum score is 0, with a lower number indicating less spasticity. | Yes |
| Cortico-spinal Tract Integrity | The investigators will measure muscle evoked potential consequent to Transcranial Magnetic Stimulation of the cortico-spinal tract to assess integrity of the cortico-spinal tract. They will also explore SCS responses when conditioned by a TMS pulse and vice-versa. | Yes |
| Sensorimotor Network Structure Integrity | The investigators will perform High-definition Diffusion Weighted Imaging to quantify Fractional Anisotropy as a measurement of axon integrity in the brain and spinal cord pre and post study. | Yes |
| Sensory motor integration: success-rate | The investigators will use the KINARM robot to quantify functional sensory acuity and sensory-motor integration. The investigators will measure 2D kinematics of the arm during different exercises where subjects will reach to defined targets with and without visual feedback. These tasks are designed to assess proprioception acuity and sensory-motor integration. Success-rate will be quantified offline. Comparison will be done per patient between Stim-on and Stim-off at different timepoints. Given the scientific nature of this task no minimal acceptable improvement is defined and data will be used to understand effects of SCS on sensorimotor integration processes. | No |
| Sensory motor integration: displacement error | The investigators will use the KINARM robot to quantify functional sensory acuity and sensory-motor integration. The investigators will measure 2D kinematics of the arm during different exercises where subjects will reach to defined targets with and without visual feedback. These tasks are designed to assess proprioception acuity and sensory-motor integration. Displacement error from the true target location will be quantified offline. Comparison will be done per patient between Stim-on and Stim-off at different timepoints. Given the scientific nature of this task no minimal acceptable improvement is defined and data will be used to understand effects of SCS on sensorimotor integration processes. | No |
| Sensorimotor Network Function | The investigators will perform resting state and motor-task functional MRI of the brain and spinal cord to quantify neural network activation at rest and during the execution of simple motor tasks. | No |
| Spinal Circuit Excitability | The investigators will measure H-reflexes of arm muscles obtained during stimulation of the peripheral nerves to quantify excitability of spinal motoneurons to stimulation of primary sensory afferents pre and post-study. Expected Result: The main scientific hypothesis is that SCS will change sensori-to-motoneuron excitability that can be measured via H-reflex responses pre and post-implant. | No |
| Motoneuron Firing Rates | The investigators will use high-density EMGs on arm muscles to calculate firing rates of single spinal motoneuron discharge during isometric maximal voluntary contractions. | No |

**Supplementary Table 2.** Time, in hours, engaged in active movement under spinal cord stimulation (SCS) ON and OFF conditions, across participants and time points.

| **time on active movement (SCS OFF/SCS ON)** | | | | | | | | |
|---|---|---|---|---|---|---|---|---|
| **time point** | **SCS01** | **SCS02** | **SCS03** | **SCS04** | **SCS05** | **SCS07** | **SCS08** | **AVERAGE** |
| pre-implant | 0.9/0 | 0/0 | 0.3/0 | 0.7/0 | 3.6/0 | 1.6/0 | 3.1/0 | **1.5/0** |
| **SCS intervention (post-implant)** | | | | | | | | |
| week 1 | 0.4/0.7 | 0.3/0.5 | 1.0/0.9 | 0.6/1.1 | 0.6/1.6 | 0.9/1.1 | 0.7/2.1 | **0.6/1.1** |
| week 2 | 0.3/0.8 | 0.4/0.6 | 0.4/1.0 | 0.6/1.3 | 1.5/2.1 | 1.2/1.5 | 1.2/2.3 | **0.8/1.4** |
| week 3 | 0.7/1.5 | 0.4/1.0 | 0.6/0.8 | 0.5/1.1 | 1.7/2.7 | 1.0/1.8 | 1.2/2.8 | **0.9/1.7** |
| week 4 | 0.4/0.4 | 0.6/1.0 | 0.5/0.9 | 0.8/1.3 | 1.2/2.0 | 0.7/1.3 | 1.1/2.2 | **0.8/1.3** |
| **TOTAL (SCS intervention)** | 1.8/3.4 | 1.6/3.0 | 2.5/3.6 | 2.5/4.8 | 4.9/8.3 | 3.8/5.7 | 4.2/9.4 | **3.1/5.5** |

**Supplementary Table 3.** Fugl-Meyer Assessment (FMA) scores for upper-extremity motor function under spinal cord stimulation (SCS) ON and OFF conditions, across participants and time points. Total scores and subcomponent scores are reported.

| Fugl-Meyer Assessment (FMA) for upper-extremity motor function | | | | | | | | |
|---|---|---|---|---|---|---|---|---|
| **time point** | **SCS01** | **SCS02** | **SCS03** | **SCS04** | **SCS05** | **SCS07** | **SCS08** | **TOTAL** |
| **PRE-IMPLANT (BASELINE)** | | | | | | | | |
| Reflexes | 4 | 4 | 4 | 4 | 4 | 4 | 4 | 4 |
| Flexor Synergy (FS) | 8 | 5 | 6 | 6 | 6 | 7 | 9 | 12 |
| Extensor Synergy (ES) | 4 | 2 | 6 | 5 | 5 | 3 | 5 | 6 |
| Movement Combining Synergy (MCS) | 3 | 0 | 3 | 2 | 0 | 2 | 3 | 6 |
| Movement Out-of-Synergy (MOS) | 1 | 0 | 1 | 0 | 1 | 0 | 1 | 6 |
| Normal Reflex Activity | 0 | 0 | 0 | 0 | 0 | 0 | 0 | 2 |
| Wrist (WRIST) | 3 | 0 | 2 | 0 | 0 | 3 | 4 | 10 |
| Hand (HAND) | 9 | 0 | 2 | 2 | 2 | 11 | 2 | 14 |
| Coordination/Speed | 4 | 4 | 4 | 4 | 2 | 4 | 4 | 6 |
| **TOTAL** | 36 | 15 | 28 | 23 | 20 | 34 | 32 | 66 |
| **WEEK 2 (SCS OFF/SCS ON)** | | | | | | | | |
| Reflexes | 4/4 | 4/4 | 4/4 | 4/4 | 4/4 | 4/4 | 4/4 | 4/4 |
| Flexor Synergy (FS) | 8/10 | 7/7 | 8/8 | 6/6 | 5/8 | 7/8 | 10/10 | 12/12 |
| Extensor Synergy (ES) | 5/5 | 3/3 | 6/6 | 4/6 | 5/5 | 3/4 | 6/6 | 6/6 |
| Movement Combining Synergy (MCS) | 3/5 | 0/0 | 2/3 | 0/3 | 1/1 | 1/3 | 3/3 | 6/6 |
| Movement Out-of-Synergy (MOS) | 1/2 | 0/0 | 1/1 | 1/2 | 1/1 | 0/1 | 1/2 | 6/6 |
| Normal Reflex Activity | 0/0 | 0/0 | 0/0 | 0/0 | 0/0 | 0/0 | 0/0 | 2/2 |
| Wrist (WRIST) | 3/7 | 0/0 | 2/5 | 1/1 | 2/2 | 2/2 | 5/5 | 10/10 |
| Hand (HAND) | 7/11 | 0/0 | 2/2 | 1/2 | 3/3 | 11/10 | 3/3 | 14/14 |
| Coordination/Speed | 4/4 | 4/4 | 4/4 | 4/4 | 3/3 | 4/4 | 4/4 | 6/6 |
| **TOTAL** | 35/48 | 18/18 | 29/33 | 21/28 | 24/27 | 32/36 | 36/37 | 66/66 |
| **WEEK 4 (SCS OFF/SCS ON)** | | | | | | | | |
| Reflexes | 4/4 | 4/4 | 4/4 | 4/4 | 4/4 | 4/4 | 4/4 | 4/4 |
| Flexor Synergy (FS) | 10/11 | 6/6 | 11/11 | 6/6 | 8/7 | 8/9 | 10/10 | 12/12 |
| Extensor Synergy (ES) | 6/6 | 3/3 | 6/6 | 6/6 | 5/5 | 3/4 | 6/6 | 6/6 |
| Movement Combining Synergy (MCS) | 5/5 | 0/0 | 3/4 | 1/2 | 1/1 | 3/3 | 4/4 | 6/6 |
| Movement Out-of-Synergy (MOS) | 2/4 | 0/0 | 1/1 | 0/2 | 1/1 | 1/1 | 2/2 | 6/6 |
| Normal Reflex Activity | 0/0 | 0/0 | 0/0 | 0/0 | 0/0 | 0/0 | 0/0 | 2/2 |
| Wrist (WRIST) | 5/6 | 0/0 | 4/3 | 0/1 | 5/5 | 2/2 | 4/4 | 10/10 |
| Hand (HAND) | 11/11 | 1/0 | 2/4 | 2/1 | 3/3 | 10/10 | 2/3 | 14/14 |
| Coordination/Speed | 4/4 | 4/4 | 4/4 | 4/4 | 3/3 | 4/4 | 4/4 | 6/6 |
| **TOTAL** | 47/51 | 18/17 | 35/37 | 23/26 | 30/29 | 35/37 | 36/37 | 66/66 |
| **FOLLOW-UP** | | | | | | | | |
| Reflexes | 4 | 4 | 4 | - | 4 | 4 | 4 | 4 |
| Flexor Synergy (FS) | 9 | 6 | 9 | - | 7 | 7 | 10 | 12 |
| Extensor Synergy (ES) | 6 | 2 | 6 | - | 6 | 5 | 6 | 6 |
| Movement Combining Synergy (MCS) | 3 | 0 | 3 | - | 1 | 3 | 4 | 6 |
| Movement Out-of-Synergy (MOS) | 4 | 0 | 1 | - | 1 | 0 | 1 | 6 |
| Normal Reflex Activity | 0 | 0 | 0 | - | 0 | 0 | 0 | 2 |
| Wrist (WRIST) | 5 | 0 | 2 | - | 5 | 3 | 4 | 10 |
| Hand (HAND) | 11 | 1 | 2 | - | 2 | 10 | 3 | 14 |
| Coordination/Speed | 4 | 4 | 4 | - | 2 | 4 | 4 | 6 |
| **TOTAL** | 46 | 17 | 31 | - | 28 | 36 | 36 | 66 |

**Supplementary Table 4.** Fugl-Meyer Assessment (FMA) scores for upper-extremity sensory function under spinal cord stimulation (SCS) ON and OFF conditions, across participants and time points. Total scores and subcomponent scores are reported.

| Fugl-Meyer Assessment (FMA) for upper-extremity sensory function | | | | | | | | |
|---|---|---|---|---|---|---|---|---|
| **time point** | **SCS01** | **SCS02** | **SCS03** | **SCS04** | **SCS05** | **SCS07** | **SCS08** | **TOTAL** |
| **PRE-IMPLANT (BASELINE)** | | | | | | | | |
| Light Touch | 4 | 2 | 2 | 1 | 3 | 4 | 4 | 4 |
| Proprioception | 7 | 0 | 6 | 2 | 6 | 7 | 8 | 8 |
| **TOTAL** | 11 | 2 | 8 | 3 | 9 | 11 | 12 | 12 |
| **WEEK 2 (SCS OFF/SCS ON)** | | | | | | | | |
| Light Touch | 4/4 | 1/1 | 2/2 | 1/1 | 3/3 | 3/3 | 4/4 | 4/4 |
| Proprioception | 5/5 | 0/0 | 3/3 | 2/0 | 6/7 | 6/7 | 8/8 | 8/8 |
| **TOTAL** | 9/9 | 1/1 | 5/5 | 3/1 | 9/10 | 9/10 | 12/12 | 12/12 |
| **WEEK 4 (SCS OFF/SCS ON)** | | | | | | | | |
| Light Touch | 4/4 | 1/1 | 2/2 | 2/0 | 3/3 | 4/3 | 4/4 | 4/4 |
| Proprioception | 7/7 | 0/0 | 5/5 | 2/1 | 6/6 | 8/8 | 8/8 | 8/8 |
| **TOTAL** | 11/11 | 1/1 | 7/7 | 4/1 | 9/9 | 12/11 | 12/12 | 12/12 |
| **FOLLOW-UP** | | | | | | | | |
| Light Touch | 4 | 2 | 2 | - | 3 | 4 | 4 | 4 |
| Proprioception | 7 | 0 | 4 | - | 6 | 8 | 8 | 8 |
| **TOTAL** | 11 | 2 | 6 | - | 9 | 12 | 12 | 12 |

**Supplementary Table 5.** Action Research Arm Test (ARAT) scores under spinal cord stimulation (SCS) ON and OFF conditions, across participants and time points. Total scores and subcomponent scores are reported.

| Action Research Arm Test (ARAT) | | | | | | | | |
|---|---|---|---|---|---|---|---|---|
| **time point** | **SCS01** | **SCS02** | **SCS03** | **SCS04** | **SCS05** | **SCS07** | **SCS08** | **TOTAL** |
| **PRE-IMPLANT (Affected/Non-affected arm)** | | | | | | | | |
| Grasp | 15/18 | 0/18 | 0/18 | 0/18 | 0/18 | 4/18 | 1/18 | 18/18 |
| Grip | 8/12 | 0/11 | 2/12 | 1/11 | 2/12 | 3/11 | 2/12 | 12/12 |
| Pinch | 0/18 | 0/18 | 0/15 | 0/15 | 0/14 | 2/18 | 0/17 | 18/18 |
| Gross Movement | 8/9 | 3/9 | 0/9 | 5/9 | 3/9 | 5/9 | 5/9 | 9/9 |
| **TOTAL** | 34/57 | 3/56 | 2/54 | 6/53 | 5/53 | 14/56 | 8/56 | 57/57 |
| **WEEK 4 - SCS OFF (Affected/Non-affected arm)** | | | | | | | | |
| Grasp | 13/18 | - | 0/18 | 0/- | 3/18 | 4/18 | 1/18 | 18/18 |
| Grip | 8/12 | - | 2/12 | 1/- | 4/12 | 3/12 | 2/12 | 12/12 |
| Pinch | 8/18 | - | 1/15 | 0/- | 0/16 | 0/16 | 0/18 | 18/18 |
| Gross Movement | 7/9 | - | 7/9 | 5/3 | 3/9 | 5/9 | 5/9 | 9/9 |
| **TOTAL** | 36/57 | - | 10/54 | 6/- | 10/55 | 12/55 | 8/57 | 57/57 |
| **WEEK 4 - SCS ON (Affected/Non-affected arm)** | | | | | | | | |
| Grasp | 18/18 | - | 0/18 | 2/- | 3/18 | 4/18 | 1/18 | 18/18 |
| Grip | 8/12 | - | 5/12 | 1/- | 4/12 | 2/11 | 2/12 | 12/12 |
| Pinch | 12/18 | - | 1/15 | 0/- | 0/16 | 0/13 | 0/18 | 18/18 |
| Gross Movement | 7/9 | - | 7/9 | 5/- | 3/9 | 5/9 | 5/9 | 9/9 |
| **TOTAL** | 45/57 | - | 13/54 | 8/- | 10/55 | 11/51 | 8/57 | 57/57 |
| **FOLLOW-UP (Affected/Non-affected arm)** | | | | | | | | |
| Grasp | 18/18 | 0/18 | 5/18 | - | 0/18 | 4/18 | 1/18 | 18/18 |
| Grip | 8/12 | 0/10 | 0/12 | - | 0/12 | 3/12 | 3/12 | 12/12 |
| Pinch | 12/18 | 0/18 | 0/15 | - | 0/14 | 0/16 | 0/18 | 18/18 |
| Gross Movement | 7/9 | 3/9 | 4/7 | - | 3/9 | 5/9 | 5/9 | 9/9 |
| **TOTAL** | 45/57 | 3/55 | 9/52 | - | 3/53 | 12/55 | 9/57 | 57/57 |

**Supplementary Table 6.** Linear correlations between agonist–antagonist muscle activation ratios and arm kinematic metrics across stimulation conditions. Myoelectric activity of the triceps brachii (TRI), brachioradialis (BR), and biceps brachii (BIC) muscles was quantified as the root mean square (RMS) of the EMG signal envelope during the reach and pull phases of a planar center-out task. Agonist–antagonist activation ratios (TRI/BR and TRI/BIC) were computed under spinal cord stimulation (SCS) ON and OFF conditions. Pearson correlation coefficients and p-values were calculated to assess the linear correlation between the percentage change in muscle activation ratios (SCS ON/SCS OFF) and average changes in arm kinematic parameters (path efficiency, deviation error, velocity peaks, and log dimensionless jerk). Statistically significant correlations ($p < 0.05$) are highlighted in bold.

| **TRI/BR (SCS ON/SCS OFF)** | | | | |
|---|---|---|---|---|
| | **reach (n = 24)** | | **pull (n = 23)** | |
| | p-value | pearson coefficient | p-value | pearson coefficient |
| path efficiency | **<0.001** | **0.644** | 0.081 | -0.302 |
| deviation error | 0.136 | 0.234 | 0.067 | -0.322 |
| velocity peaks | **<0.001** | **0.609** | 0.365 | -0.076 |
| log dimensionless jerk | **0.004** | **0.530** | 0.186 | -0.195 |

| **TRI/BIC (SCS ON/SCS OFF)** | | | | |
|---|---|---|---|---|
| | **reach (n = 24)** | | **pull (n = 23)** | |
| | p-value | pearson coefficient | p-value | pearson coefficient |
| path efficiency | **0.013** | **0.452** | **0.029** | **-0.399** |
| deviation error | 0.786 | 0.264 | 0.202 | -0.183 |
| velocity peaks | **<0.001** | **0.613** | **<0.001** | **-0.649** |
| log dimensionless jerk | **0.006** | **0.502** | **<0.001** | **-0.612** |

**FIGURES**

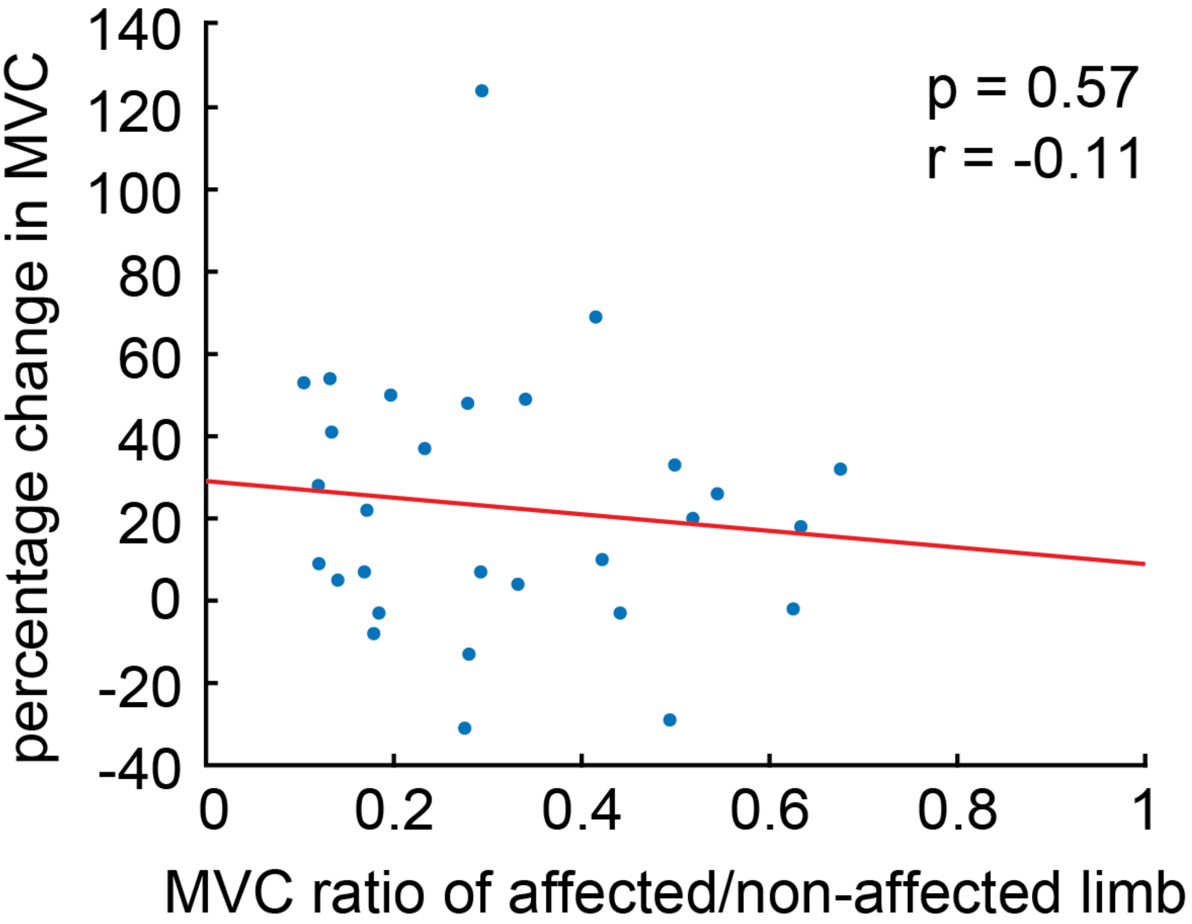


**Supplementary Fig. 1 | Correlation between the percentage change in Maximum Voluntary Contraction (MVC) strength under spinal cord stimulation (SCS) conditions and impairment.**
Isometric MVC during SCS ON and OFF conditions were used to assess strength of shoulder flexion, shoulder extension, elbow flexion, elbow extension and grip for all participants, except SCS02 since the MVC of the non-affected limb was not measured for this participant. Across all isolated movements, the ratio between the MVC force produced by the affected and non-affected limb showed no correlation with the percentage change in MVC strength.

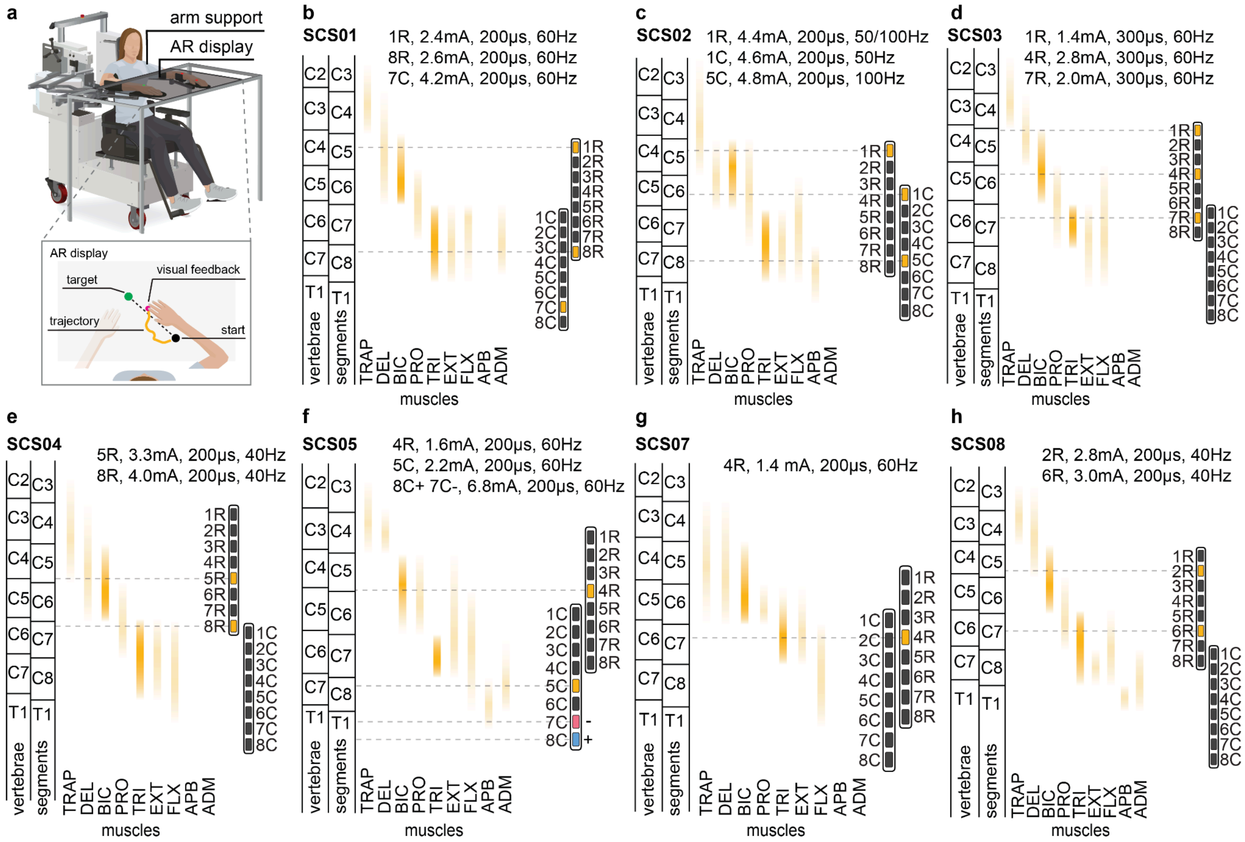


**Supplementary Fig. 2 | Spinal segment maps and spinal cord stimulation (SCS) configuration during arm dexterity reach-and-pull task.**

**a.** Experimental setup for reach-and-pull task using an exoskeleton robot (KINARM) for anti-gravity arm support. **b-h.** Spinal segment maps indicating the spinal levels and motor neuron pools targeted by the electrodes used for each participant (SCS01 - SCS08) during the task. Abbreviations: R, contacts in the most rostral lead; C, contacts in the most caudal lead; TRAP, trapezius; DEL, deltoids; BIC, biceps brachii; PRO, pronator; TRI, triceps brachii; EXT, wrist extensors; FLX, wrist flexors; APB, abductor pollicis brevis; ADM, abductor digiti minimi.

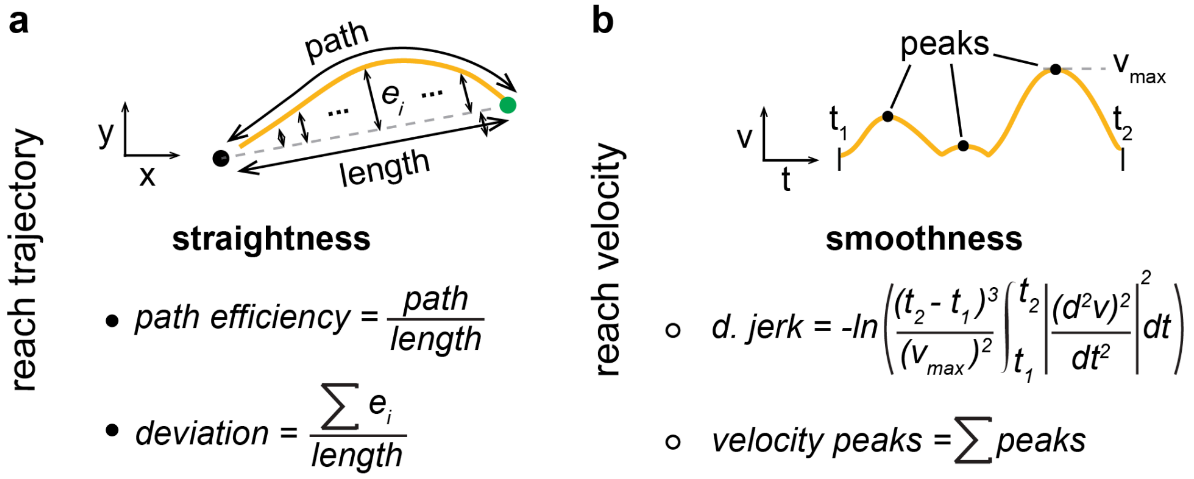


**Supplementary Fig. 3 | Arm kinematic metrics**.
Reach and pull trajectories were quantified using **a.** straightness metrics (path efficiency ratio and deviation error) and **b.** smoothness metrics (log dimensionless jerk and number of velocity peaks).

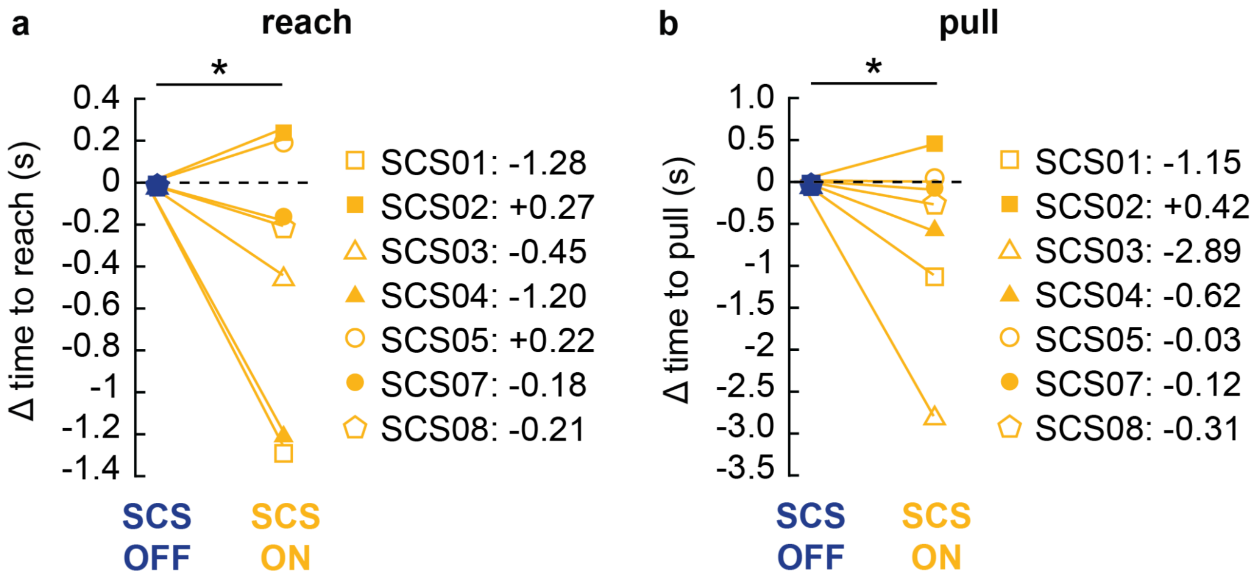


**Supplementary Fig. 4 | Change in duration of reach and pull phases under spinal cord stimulation (SCS)**

Significant decrease in duration, measured in seconds (s), of **a.** reach (average -0.41s) and **b.** pull (average -0.67s) phases under SCS ON compared to SCS OFF condition across all participants. Movement durations for both SCS ON and OFF conditions were subtracted by the duration of the SCS OFF condition.

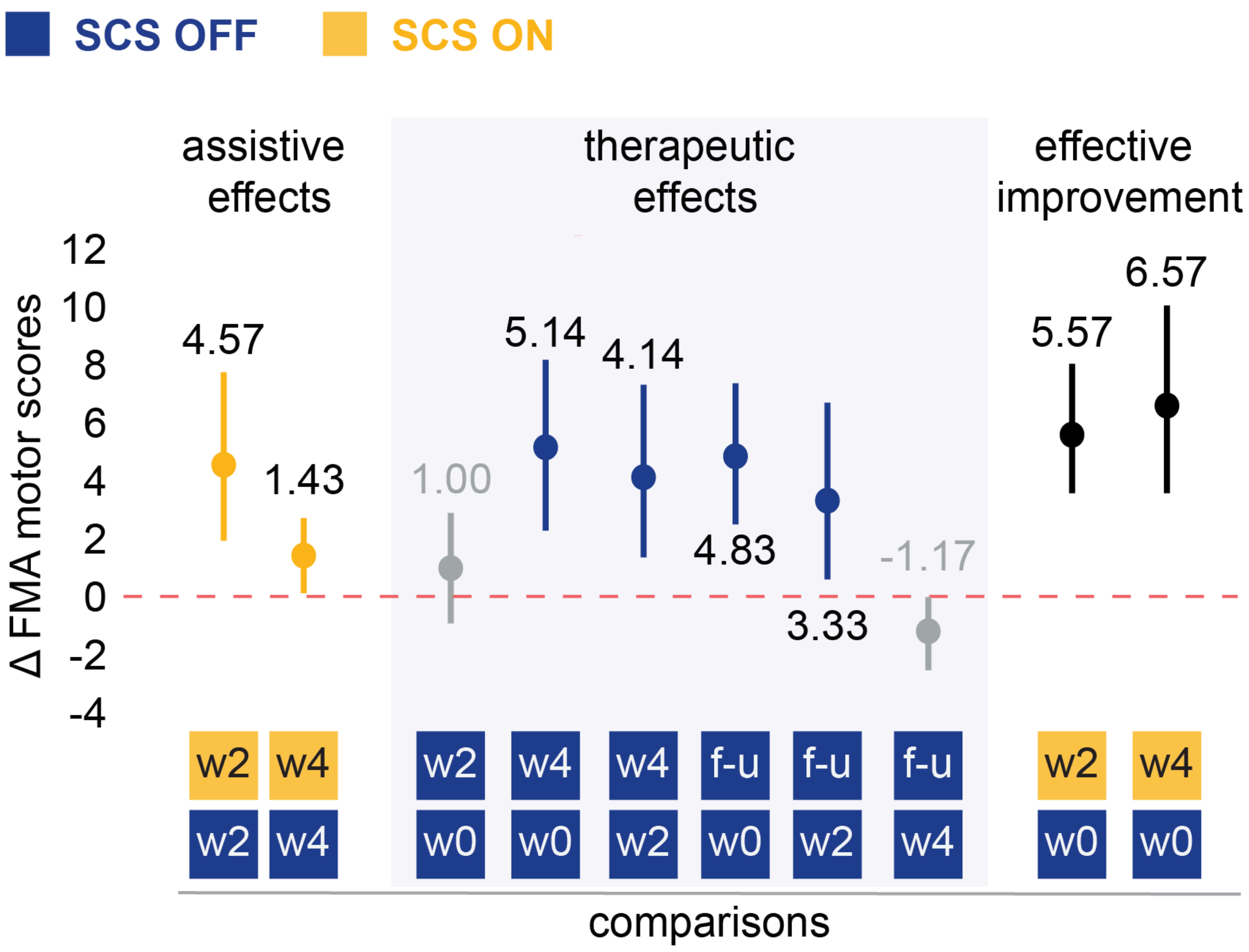


**Supplementary Fig. 5 | Average change in the Fugl-Meyer Assessment (FMA) scores for upper-extremity motor function across spinal cord stimulation (SCS) ON and OFF conditions.** Assistive effects of SCS were measured as the difference in FMA scores between SCS ON and OFF conditions at week 2 (w2) and week 4 (w4). Therapeutic effects of SCS were measured as the difference in FMA scores between time points (pre-implant, w0; week 2, w2; week 4, w4; follow-up, f-u) under SCS OFF condition. Effective improvement was measured as the difference in FMA scores between SCS OFF at pre-implant (w0) and SCS ON at week 2 (w2) and week 4 (w4).
